# Comparative effectiveness of preferred pharmacological treatment options for bipolar disorder among people with opioid use disorder in British Columbia and Ontario, Canada: protocol for parallel population-based target trial emulations

**DOI:** 10.64898/2026.04.02.26350000

**Authors:** Md. Belal Hossain, Ruyu Yan, Kristen A Morin, Martin Rotenberg, Angela Russolillo, Marco Solmi, Tasneem Lalva, David C Marsh, Bohdan Nosyk

**Affiliations:** Centre for Advancing Health Outcomes, Vancouver, British Columbia, Canada; Faculty of Health Sciences, Simon Fraser University, Burnaby, British Columbia, Canada; Northern Ontario School of Medicine University, Sudbury, Ontario, Canada; Health Sciences North Research Institute, Sudbury, Ontario, Canada; Department of Psychiatry, University of Toronto, Toronto, Ontario, Canada; Centre for Addiction and Mental Health, Toronto, Ontario, Canada; School of Nursing, University of British Columbia, Vancouver, British Columbia, Canada; SCIENCES lab, Department of Psychiatry, Faculty of Medicine, University of Ottawa, Ontario, Canada; Department of Mental Health, The Ottawa Hospital, Ottawa, Ontario, Canada; Ottawa Hospital Research Institute (OHRI), Ottawa, Ontario, Canada; Department of Child and Adolescent Psychiatry, Charite Universitaetsmedizin, Berlin, Germany

## Abstract

**Introduction:** People with bipolar disorder (BD) and concurrent opioid use disorder (OUD) experience more severe clinical outcomes, including higher mortality, treatment complexity, and worse psychiatric symptoms, yet they are underserved due to a lack of tailored clinical guidelines and limited supporting research on competing treatment options. While pharmacological treatments for BD are well-established, their use varies widely across settings, and their effectiveness in individuals with co-occurring OUD is unclear. We propose parallel population-based studies to emulate randomized controlled trials to assess the comparative effectiveness of pharmacological treatment options for BD among people with OUD in British Columbia and Ontario, Canada, 2010-2023.

**Methods and analysis:** We propose emulating a series of parallel target trials using linked population-level health administrative data for all individuals aged 18 years or older diagnosed with both BD and OUD and who initiated treatments for BD between 1 January 2010 and 31 December 2023. All analyses will be conducted in parallel in British Columbia and Ontario. We propose a series of four successive target trial emulations, comparing (i) lithium versus non-antipsychotic mood stabilizers such as divalproex, lamotrigine, and valproic acid; (ii) lithium versus 2^nd^ generation antipsychotics with mood stabilizing properties such as risperidone, olanzapine, aripiprazole, and quetiapine; (iii) lithium versus combination treatments such as lithium and divalproex, lithium and olanzapine, lithium and aripiprazole, lithium and quetiapine, divalproex and olanzapine, and olanzapine and quetiapine; (iv) lithium and valproate (LATVAL) versus lithium and olanzapine, lithium and aripiprazole, lithium and quetiapine, divalproex and olanzapine, and olanzapine and quetiapine. Incident user and prevalent new user analyses are planned for proposed target trials (i)-(iv), pending sufficient data. Stratified analyses will be conducted for BD-I, manic and depressive phases of BD illness. We propose an initiator analysis (intention-to-treat, conditional on medication dispensation) to determine the effectiveness of the treatments and per-protocol analyses to determine the efficacy of the treatments after dealing with treatment switching and recommended dose adjustment. The outcomes will include psychiatric acute-care visits (hospitalizations and emergency department visits), BD treatment discontinuation and all-cause mortality. Subgroup and sensitivity analyses, including cohort and study timeline restrictions, eligibility criteria modifications, and outcome reclassifications, are proposed to assess the robustness of our results. Executing analyses in parallel across settings using a co-developed protocol will allow us to evaluate the replicability of findings.

**Ethics and dissemination:** The protocol, cohort creation, and analysis plan have been classified and approved as a quality improvement initiative by the Providence Health Care Research Ethics Board and the Simon Fraser University Office of Research Ethics. Results will be disseminated to local advocacy groups, clinical groups and decision-makers, national and international clinical guideline developers, presented at international conferences, and published in peer-reviewed journals.

**Strengths and limitations of this study:**

- A target trial using linked health administrative data will be used to determine the comparative effectiveness of preferred pharmacological treatment options for bipolar disorder among people with opioid use disorder, as observed in clinical practice in British Columbia and Ontario.
- Inverse probability weighting with investigator-specified covariates will be used to address measured confounding, while high-dimensional propensity score and instrumental variable analysis will be used to address potential unmeasured confounding.
- Potential confounding bias by severity and other threats to validity will be assessed via a range of sensitivity and subgroup analyses.
- Protocol co-development and parallel population-level analyses will address limitations in data capture in each setting and allow direct evaluation of the replicability of findings.

## Introduction

### Bipolar Disorder: Definition, epidemiology and recommended treatment

Bipolar disorder (BD) is a chronic mental disorder characterized by recurrent mood episodes, including manic/hypomanic and depressed periods and subsyndromal mood symptoms.^1^ BD is one of the major contributors to disability, resulting in cognitive and functional challenges and an increased risk of mortality, especially through suicide.^2^ The global prevalence of BD is relatively low, 2.1% to 2.8% in the general population,^3^ but contributes to substantial disease burden, criminal justice involvement, and high societal costs.^4–8^

The standard of care for the majority of individuals with BD includes pharmacological treatment.^9^ Pharmacological maintenance treatment is needed for almost all individuals with BD to alleviate symptoms, prevent relapse and restore functioning.^8^ Lithium, divalproex, and several second-generation antipsychotics, including risperidone and quetiapine, are among the most common first-line treatments for managing BD.^9–15^ A meta-analysis of 39 randomized controlled trials (RCTs) reported that lithium, risperidone, and quetiapine resulted in greater efficacy compared with placebo.^16^ Lithium or valproate (LATVAL) plus an antipsychotic is also the first-line pharmacological treatment option for BD with severe manic or mixed BD episodes.^11,17^ For patients experiencing hypomania, monotherapy with lithium, valproate, or an antipsychotic is recommended.^9,11^ An overview of systematic reviews also confirmed the efficacy of valproate across phases, showing benefits in acute mania, bipolar depression, and maintenance treatment.^18^ Moreover, lamotrigine, commonly used for maintenance therapy, has also been found to reduce the recurrence of manic episodes. Second-generation antipsychotics such as olanzapine, risperidone, ziprasidone, and aripiprazole are also recommended for treating acute manic or mixed episodes in BD.^19^ Second-generation antipsychotics are generally preferred over first-generation antipsychotics due to their superior safety profile, with fewer extrapyramidal side effects reported.^20^ There is also consistent evidence in the literature for the effectiveness of pharmacologic treatment options for BD with combination therapy demonstrating superior effectiveness. A meta-analysis including 113 RCTs showed that lithium, valproate, and several second-generation antipsychotics are superior to placebo for acute mania and are generally more effective than monotherapy.^21^

### Prevalence and treatment of concurrent BD and OUD

It is estimated that 50-90% of people with opioid use disorder (OUD) have a concurrent mental disorder such as major depression, schizophrenia or BD.^22,23^ A systematic review of 34 observational studies reported a prevalence of BD among people with OUD of 8.7% (95% confidence interval [CI] 6.7%-10.7%).^24^ Concurrent OUD challenges the effective treatment of BD in several ways. First, both BD and OUD involve dopaminergic dysregulation, and chronic opioid use may desensitize or alter dopamine signalling.^25,26^ Both are also linked to dysregulation of the hypothalamic-pituitary-adrenal (HPA) axis, which governs the stress response. Both of these factors may reduce the effectiveness of mood stabilizers such as lithium or atypical antipsychotics,^27–29^ necessitating different dosing strategies. Second, since OUD is associated with alterations in neuroplasticity and may cause structural changes in the prefrontal cortex and amygdala, treatments, such as lithium, that modulate multiple pathways to facilitate neural plasticity, may be less effective.^30,31^ Anti-inflammatory adjuncts or immune-modulating strategies may be more useful for those with concurrent BD and OUD, given that both conditions are associated with neuroinflammation.^32,33^ Third, medications for BD and OUD may have potential drug-drug interactions, increasing the risk of adverse effects such as central nervous system depression, QT prolongation, serotonin syndrome, and liver dysfunction.^34,35^ Fourth, OAT may complicate or mask depressive symptoms in BD. Serotonergic dysregulation is strongly implicated in mood disorders, with reduced serotonin levels commonly associated with depression.^36^ Both methadone and buprenorphine/naloxone have been shown to increase serotonin activity, which may alter mood expression and complicate clinical assessment.^35,37^ Finally, BD symptoms are often masked in people with OUD, contributing to an average 10-year delay in BD diagnosis, regardless of concurrent drug use. Drug use may further mirror BD symptoms, leading to additional delays in diagnosis.^38^

People with concurrent OUD and BD are more likely to be prescribed multiple psychiatric medications^39^ and are less likely to receive the standard of care for OUD,^40^ increasing the likelihood of poor outcomes. Similarly, people with BD report significantly higher rates of substance use during opioid agonist therapy (OAT) and worse physical and psychiatric symptoms when compared to those with depression or anxiety disorders.^41–43^ Cognitive deficits and poor treatment retention observed in OUD may also partially explain the clinical complexity and poorer outcomes seen in people with co-occurring BD and OUD.^44,45^ As a result, OUD exacerbates the course of BD by worsening individual episodes and contributing to a more severe overall trajectory, including earlier onset, more frequent and mixed episodes, and slower symptom remission.^46^ Concurrent OUD and BD increase the severity of each condition, complicate treatment and may compromise adherence to both forms of treatment.^24^ The elevated risk of mortality among people with OUD is most pronounced among those with concurrent BD, whose mortality risk is 40% higher than those with no mental disorder.^23^

Among people with OUD, management of BD medications often follows addiction-focused treatment strategies, with psychiatric care primarily oriented towards stabilizing substance use. However, much of the existing literature examines BD and OUD separately, despite the high prevalence of their co-occurrence.^47–49^ Both Canadian and US guidelines provide no evidence-based recommendations specific to individuals with OUD and BD, regardless of their receipt of OAT. Canadian guidelines cite a paucity of evidence regarding BD management in this population and therefore offer no explicit guidance for treating individuals with OUD^9,50^ (**Appendix Table 1**). Consistent with this gap, a systematic review indicated that evidence supporting pharmacological interventions for co-occurring BD and substance use disorder remains weak; most relevant studies identified were small, uncontrolled and methodologically limited.^51^

**Table 1.** Administrative databases will be used to determine the comparative effectiveness of treatment options for bipolar disorder among people with opioid use disorder in British Columbia (BC) and Ontario, Canada.

| BC data source | Ontario data source | Time period | Original purpose and type of data |
| --- | --- | --- | --- |
| Client Roster (CR)<sup>86</sup> | Registered Persons Database (RPDB) | CR: 1986 – present<br>RPDB: 1991 – present | Administrative registry used to identify all provincially insured residents and capture demographic (e.g., age, sex) and geographic (e.g., postal code, health authority) information. |
| PharmaNet<sup>87</sup> | Ontario Drug Benefit (ODB) database and Narcotics Monitoring System (NMS) | PharmaNet: 1996 – present<br>ODB: 1991 – present<br>NMS: 2014 – present | PharmaNet captures drug dispensations in BC, while ODB database captures dispensations for public drug coverage in Ontario (~89% of the OUD population). In Ontario, NMS database captures data on dispensed prescriptions for narcotics, controlled substances and other monitored drugs, which is already captured by PharmaNet in BC. |
| Vital Statistics<sup>88</sup> | Vital Statistics | BC: 1986 – present<br>Ontario: 1990 – present | Capturing all deaths in the province. |
| Medical Services Plan (MSP)<sup>89</sup> | Ontario Health Insurance Plan (OHIP) Claims Database | MSP: 1985 – present<br>OHIP: 1991 – present | Capturing physician billing and outpatient service records (diagnoses, procedures, service dates). |
| Discharge Abstract Database (DAD)<sup>90</sup> | DAD | BC: 1985 – present<br>Ontario: 1988 – present | Capturing inpatient hospitalizations and day surgeries (diagnostic/procedure codes). |
| National Ambulatory Care Reporting System database (NACRS)<sup>91</sup> | NACRS | BC: 2000 – present<br>Ontario: 2000 – present | Capturing emergency department visits and ambulatory procedures. |
| Perinatal Database<sup>92</sup> | MOMBABY | BC: 1992 – present<br>Ontario: 2002 – present | Capturing maternal and infant health data for all births (e.g., prenatal care, labour, neonatal outcomes). |
| Social Development and Poverty Reduction (SDPR) Database<sup>93</sup> | - | SDPR: 1996 – present | Capturing social assistance receipt. |
| BC Provincial Corrections<sup>94</sup> | - | BC: 2010 – present | Capturing administrative data on incarcerations within provincial correctional facilities. |

Abbreviations – BC: British Columbia; CR: Client Roster; DAD: Discharge Abstract Database; MSP: Medical Services Plan; NACRS: National Ambulatory Care Reporting System; NMS: Narcotics Monitoring System; ODB: Ontario Drug Benefit; OHIP: Ontario Health Insurance Plan; OUD: opioid use disorder; RPDB: Registered Persons Database; SDPR: Social Development and Poverty Reduction.

In general, existing guidelines recommend the initiation of lithium or lamotrigine as a first-line pharmacological treatment for BD among people with OUD. Gradual titration of lamotrigine is needed and may be problematic in individuals who are inclined to take excess medication doses, and drug-drug interactions may alter lamotrigine levels and increase the risk of adverse outcomes, such as Stevens-Johnson syndrome.^52^ Antidepressant monotherapy is not recommended due to concerns about precipitating a mixed or manic episode or contributing to the development of rapid cycling.^53^ For treatment-resistant or severely impaired cases, augmentation with an antidepressant may be considered only as a last resort, and should be accompanied by a mood stabilizer such as lithium to mitigate the risk of mania.^8,9^ Electroconvulsive therapy may also serve as an adjunctive option for individuals who do not respond adequately to pharmacological treatment.^54^

The evidence on the effectiveness of treatment options for BD among people with OUD is extremely limited. The level of evidence for BD treatment options in this population is low due to a paucity of data, the complexity of study designs (given that many individuals use more than one substance), and inconsistency in the outcome variables used across studies, which hinders direct comparison of results. Importantly, prior studies are typically industry-led, exclude people with OUD, and are often characterized by small and potentially unrepresentative sample sizes.^55^ Managing BD with pharmacotherapies is further complicated by the need to tailor treatment to symptom severity, acuity (particularly during manic episodes), and episode type (depressive or manic).^9,55,56^ As such, treatment options must be evaluated for both efficacy and tolerability by prescribers. These challenges to the generalizability of the available evidence have resulted in calls for large administrative data studies to inform clinical practice.^55^

### Equipoise and potential value of the target trial framework using health administrative data

RCTs are the ‘gold standard’ for assessing BD treatment effects among people with OUD because of their ability to balance measured and unmeasured confounding and eliminate the possibility of selection bias, thereby achieving exchangeability between treatment and control groups. However, a systematic review highlighted that 55% to 96% of individuals with BD would not meet the eligibility criteria for medication-focused RCTs based on common inclusion/exclusion criteria.^57^ Additionally, retaining people with concurrent substance use disorders and serious mental illness in RCTs can be challenging, and there are very few high-quality RCTs in this population.^58^ Moreover, RCTs have important logistical challenges and limitations regarding external validity. RCTs investigating treatment outcomes in both BD and OUD commonly exclude individuals with other comorbid conditions.^59,60^ For example, the majority of RCTs comparing BD treatment exclude people with OUD (**Appendix Table 2**). In contrast, studies compared different mood stabilizers such as lithium and valproate, antipsychotics such as quetiapine, risperidone, olanzapine, and aripiprazole, along with newer or adjunctive options.^61,62,71–79,63–70^

**Table 2.** Key components of the protocol of the target trial of the pharmacological treatment options for bipolar disorder among people with opioid use disorder.

| Component | Target trial protocol | Emulated trial using observational data |
| --- | --- | --- |
| Eligibility criteria | Individuals aged $\geq 18$ years with opioid use disorder who are diagnosed with bipolar disorder (BD), with the following eligibility criteria: no schizophrenia, no kidney or liver toxicity, not receiving cancer treatment or palliative care, non-pregnant or not planning for pregnancy, and not incarcerated. | Same eligibility criteria in BC. Same eligibility criteria in Ontario, except for incarceration status.<br>Incident user design: No prior experience of receiving treatments following BD dating back to January 1, 1996 in British Columbia and January 1, 2004 in Ontario.<br>Prevalent new user design: No history of receiving treatments following BD within the past 30 days.<br>Separate analyses for BD-I, manic and depressive phases. |
| Time zero | Date of initial BD treatment assignment. | Date of initial dispensation of psychotropic medication following first indication of BD. |
| Treatment strategies | Trail 1: Lithium versus non-antipsychotic mood stabilizers such as divalproex, lamotrigine, and valproic acid;<br>Trail 2: Lithium versus 2<sup>nd</sup> generation antipsychotics with mood stabilizing properties such as risperidone, olanzapine, aripiprazole, and quetiapine;<br>Trail 3: Lithium versus combination treatments such as lithium and divalproex, lithium and olanzapine, lithium and | Same. |
|  | aripiprazole, lithium and quetiapine, divalproex and olanzapine, and olanzapine and quetiapine;<br>Trail 4: Lithium and valproate (LATVAL) versus lithium and olanzapine, lithium and aripiprazole, lithium and quetiapine, divalproex and olanzapine, and olanzapine and quetiapine. |  |
| Treatment assignment | Individuals are randomly assigned to the treatment strategies at time zero. | Treatment strategies are assigned via propensity score weighting. |
| Outcomes | Psychiatric acute-care visits, BD treatment discontinuation and all-cause mortality. The secondary outcomes are all-cause and cause-specific acute-care visits, all-cause and cause-specific hospitalizations, all-cause and cause-specific emergency department visits, BD treatment switching, on-treatment mortality, suicide mortality, positive symptoms scale, negative symptoms scale, and mania scale. | Same. Scale outcomes are applicable only to Ontario since data are not available in BC. Acute care visits will be observed via hospitalizations or emergency department visits. Mortality information will be observed via linkage to vital statistics records. |
| Follow-up | Follow-up begins at time zero and ends in outcome of interest, death, loss to follow-up or study end. | Same. |
| Causal contrast | Intent-to-treat (ITT), per-protocol (PP). | Initiator (ITT conditional on medication dispensation), PP. |
| Analysis plan | ITT: Kaplan-Meier survival curve and Cox-PH proportional hazards model.<br>PP: Adjusted Kaplan-Meier survival curve and marginal structural modelling via inverse probability of adherence weighting approach to deal with treatment switching and dose adjustment. | Initiator: Adjusted Kaplan-Meier survival curve and Cox proportional hazards model via propensity score weighting.<br>PP: Same. |

Abbreviations – BD: bipolar disorder; ITT: Intent-to-treat; PP: per-protocol.

In Canada, there is notable variation in how BD is managed among individuals with OUD. A retrospective study of 136,628 individuals with BD in Alberta, Canada, showed a notable shift in prescribing for BD between 2008 and 2021, with antidepressants remaining the most common, while the use of lithium, divalproex, and carbamazepine declined, and second-generation antipsychotics, particularly quetiapine, became increasingly common.^80^ Provinces also differ in prescriber training, management strategies, laboratory monitoring policies, and delivery methods.^81^ In the absence of evidence-based guidelines specifically addressing BD treatment with comorbid OUD, clinicians manage complex concurrent disorder using different approaches, and the extent of coordination between mental health and substance use care varies widely across Canada despite integrated care being recommended as a standard.^82,83^

### Study objective

In the present study, we propose to emulate a ‘target trial’ to assess the comparative effectiveness of the most common pharmacological treatments for BD among individuals with OUD.^84^ People with concurrent OUD and BD are unlikely to participate in conventional RCTs. Therefore, there is a need for our proposed target trial at the population level. Our objective will be to replicate the structure of RCTs using linked health administrative databases from British Columbia (BC), Canada (2010-2022), and Ontario, Canada (2014-2023). This entails establishing eligibility criteria according to data available at time zero (treatment initiation), carefully defining treatment strategies, and determining whether an ‘intent-to-treat’ or ‘per-protocol’ analysis is most appropriate. Doing so with data from two of the three most populous Canadian provinces provides an opportunity to evaluate the replicability of our findings and contrast results across settings.^85^ All analyses will be conducted in parallel in BC and Ontario.

## Methods

### Study setting and data sources

This study will employ linked population-level administrative databases in two Canadian provinces that encompass 80% of all OAT clients in Canada. Given restrictions on their interprovincial portability, all analyses will be executed in parallel in BC and Ontario. While individual-level data cannot be shared across provinces, the core study design will be consistent across settings and analytic code, and definitions (e.g., inclusion/exclusion criteria), covariate lists, and drug identification numbers will be harmonized and shared to ensure consistency in methodology across provinces.

Although there is a high degree of equivalency in data capture between the two provinces (**Table 1**), we highlight five key differences that will affect interpretation and thus have informed our study design; 1) Period of data capture: 2010-2022 in BC and 2014-2023 in Ontario; 2) ICD-9/10 codes: all health administrative databases in BC include ICD-9/10 codes up to the 5th digit, whereas Ontario databases include up to the 3rd digit in most cases. As such, there is a possibility of misclassification in medical conditions; 3) Drug dispensation databases: PharmaNet captures all BC residents, whereas the Ontario Drug Benefit (ODB) database captures Ontario residents eligible for public drug coverage, including individuals whose drug costs surpass their income. The ODB database includes prescription medication claims for people who receive drug benefits covered by the provincial government, with an estimated error rate of 0.7%. Eligibility for coverage includes individuals who are unemployed, have a disability, receive home care services, reside in long-term care, have high prescription drug costs relative to household income, or are under age 25 with ‘OHIP+’ coverage, which has been available since January 1, 2018.

Since we will link the data from the Narcotics Monitoring System (NMS), which collects data on narcotic prescriptions regardless of payment mechanism, data capture regarding OAT in Ontario will be comparable to BC. However, other prescription data related to BD will only include prescription medication claims for people who receive drug benefits covered by the provincial government; 4) Clinical scales: unique to Ontario is the Ontario Mental Health Reporting System (OMHRS), a Canadian Institute for Health Information-maintained dataset that captures all adult inpatient mental health admissions in the province. OMHRS uses the Resident Assessment Instrument-Mental Health (RAI-MH) version 2.0 as its standardized clinical assessment instrument, generating a range of validated clinical scales comparable to those commonly used in RCTs. These measures will serve as secondary outcomes for evaluating the subset of the Ontario cohort with psychiatric hospitalizations; 5) Provincial correction data: we will use incarceration as both an exclusion criterion and as a covariate, since data are available only in BC. Differences in data capture across settings will thus provide opportunities for inference beyond what is possible in either setting alone, and also provide a direct means of assessing the replicability of findings across settings.

### Study population

The study will include individuals with OUD aged 18-105 years and diagnosed with BD who receive at least one BD treatment dispensation following BD diagnosis. The case definition of OUD, BD, and BD phases is presented in **Appendix Table 3**. Those who are diagnosed with schizophrenia at time zero will be excluded. We will also exclude individuals with kidney or liver toxicity, since these individuals need expert guidance on treatment management because of impaired drug metabolism.^95^ Individuals with a cancer treatment or receiving palliative care will be excluded since cancer treatments and palliative care often involve complex medication regimens, which often require modifying the profile of prescribed drugs.^96,97^ Pregnant women will also be excluded since the pharmacokinetics of BD treatments are altered during pregnancy, and pregnant women need expert guidance on BD treatment management to avoid fetal risks.^98^ In BC, we will further exclude those who are incarcerated at time zero since these individuals often experience additional psychosocial distress, potentially influencing their eligibility to receive different BD treatments.^99,100^ To align the analyses for both provinces, we will perform a sensitivity analysis by including individuals in BC regardless of their incarceration status.

**Table 3.**
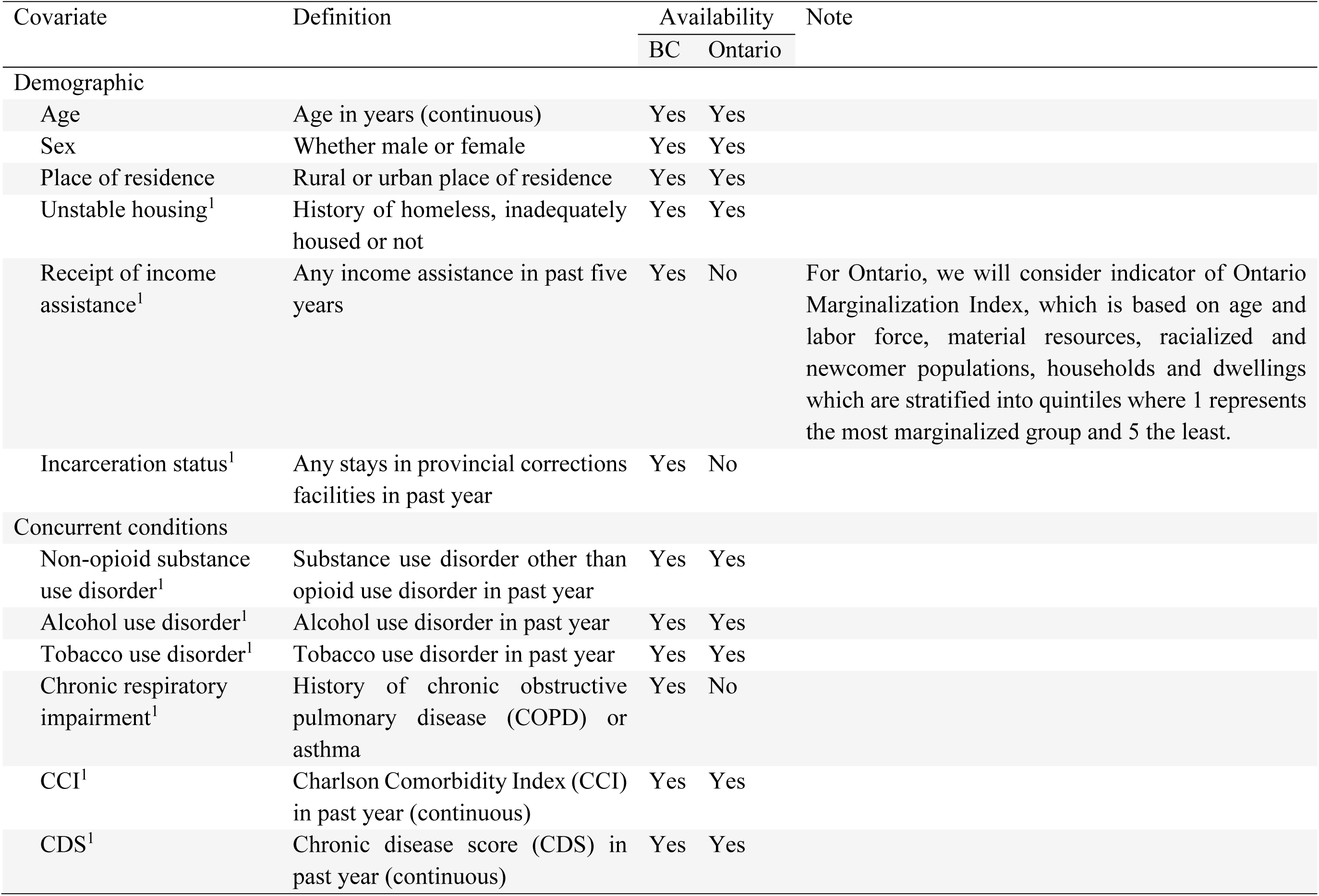
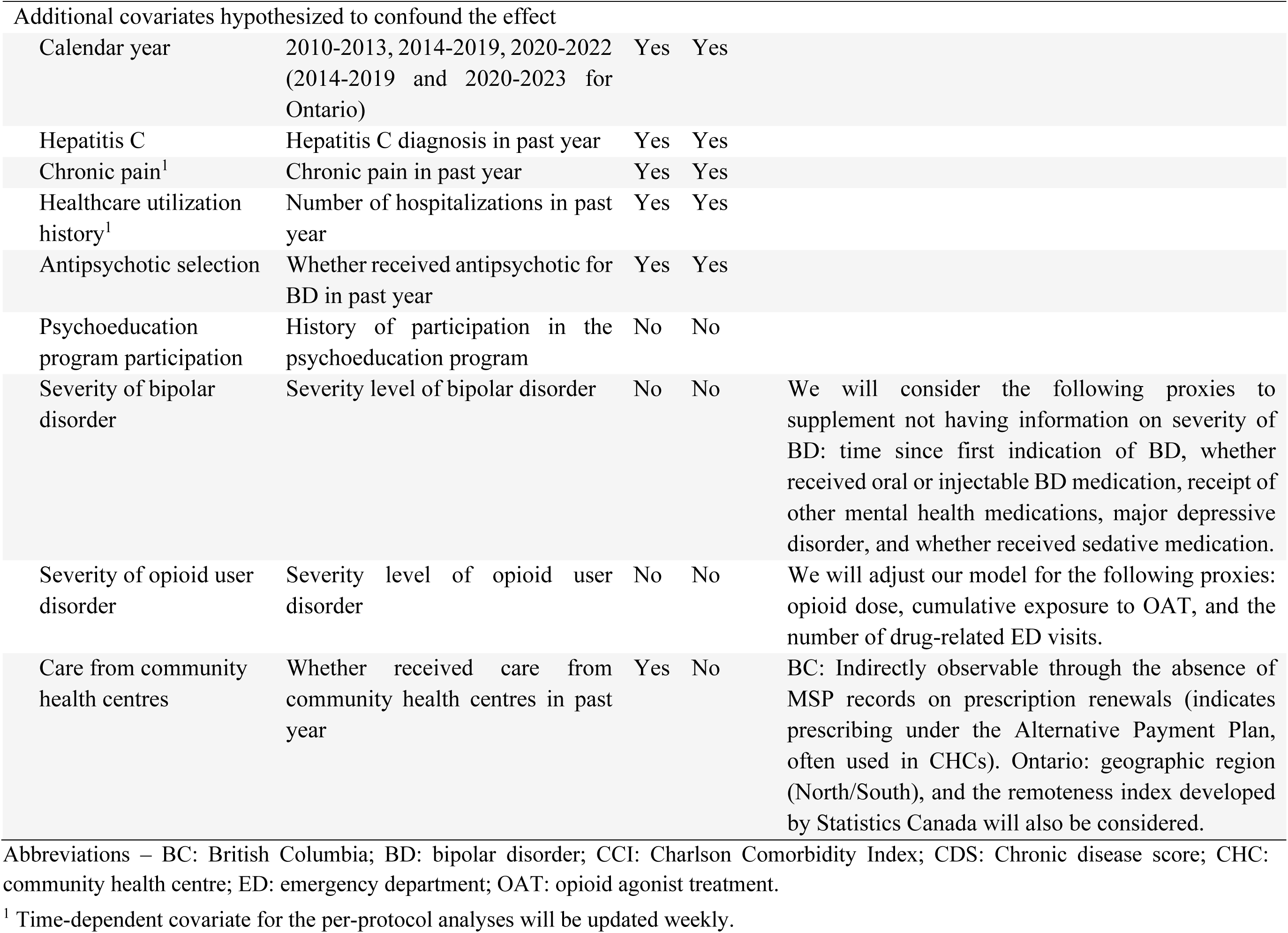
Potential confounding variables influencing the relationship of treatment options for bipolar disorder with treatment acute-care visits and all-cause mortality among people with opioid use disorder and bipolar disorder.

### Study design

**Table 2** summarizes the key components of the proposed target trial emulation. We will execute both incident-user and prevalent new-user study designs to ensure that the clinical experience of those accessing treatment across successive attempts is captured in our analyses. Incident users include individuals without a history of pharmacological treatments for BD dating back to 1 January 1996 in BC and 1 January 2004 in Ontario. To define prevalent new users, we will use a 30-day washout period, entailing no dispensations of pharmacological treatments for BD in the 30 days prior to treatment episode initiation.^101^

### Study follow-up

The study follow-up will start from time zero (first dispensation of psychotropic medication following BD diagnosis) and will end in outcome of interest (defined below), death, loss to follow-up, or study end (31 December 2022 in BC or 31 December 2023 in Ontario).

### Treatment strategies

The treatment of interest is the receipt of non-antipsychotic mood stabilizers, as well as antipsychotics with mood-stabilizing properties for BD. Four distinct trials, each with a number of arms, will be considered. The first set of treatment strategies includes lithium versus other mood stabilizers such as divalproex, lamotrigine, and valproic acid. The second set of treatment strategies includes lithium versus second-generation antipsychotics with mood-stabilizing properties, such as risperidone, olanzapine, aripiprazole, and quetiapine. The third set includes lithium versus combination treatments such as lithium and divalproex, lithium and olanzapine, lithium and aripiprazole, lithium and quetiapine, divalproex and olanzapine, and olanzapine and quetiapine. The fourth set includes LATVAL versus lithium and olanzapine, lithium and aripiprazole, lithium and quetiapine, divalproex and olanzapine, and olanzapine and quetiapine, pending sufficient data. The details of the treatments of interest are presented in **Appendix Table 4**. Pending sufficient data, we will conduct stratified analyses to compare the BD treatment strategies for any BD, and BD-I, manic and depressive phases.

**Table 4.**
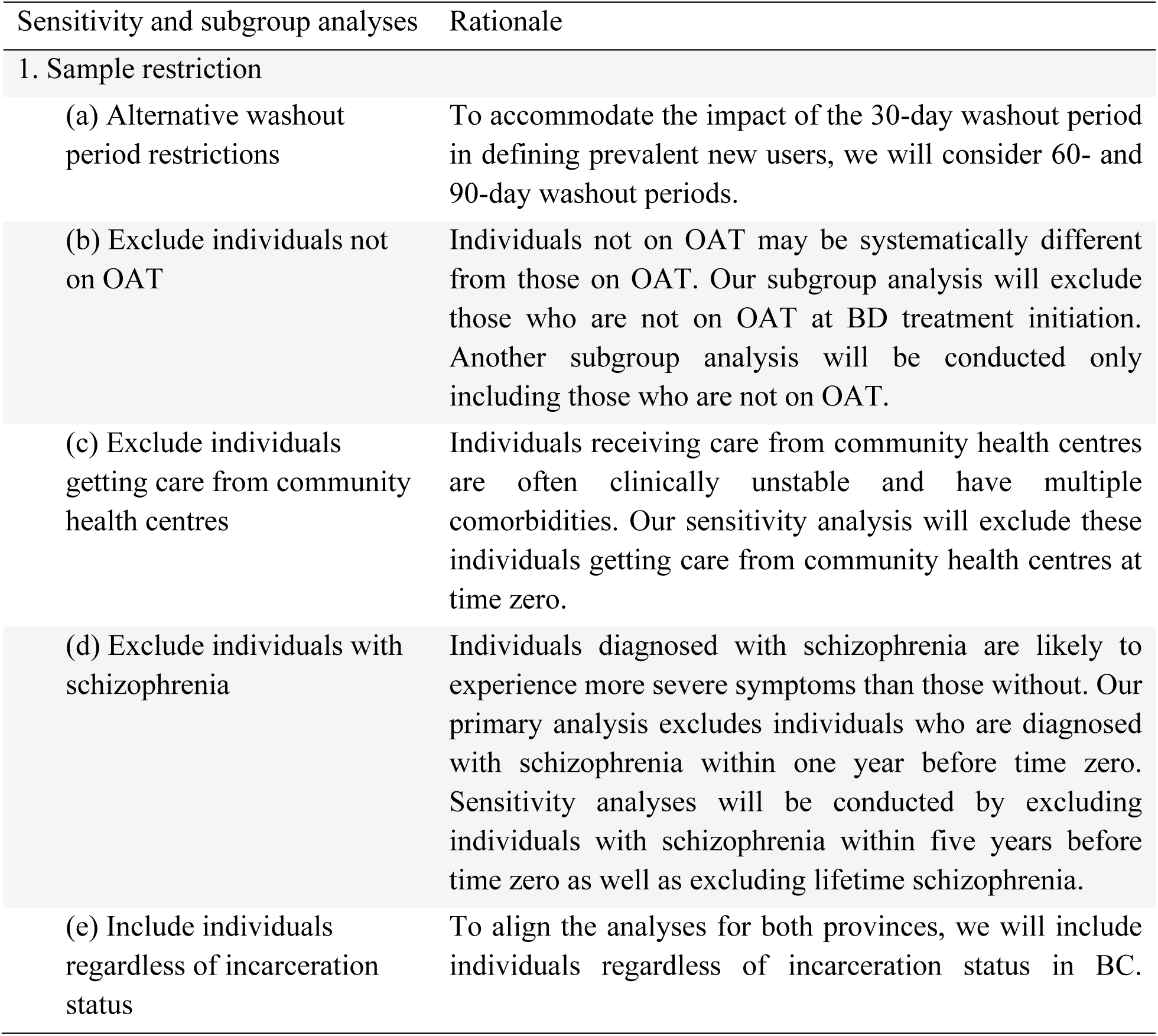
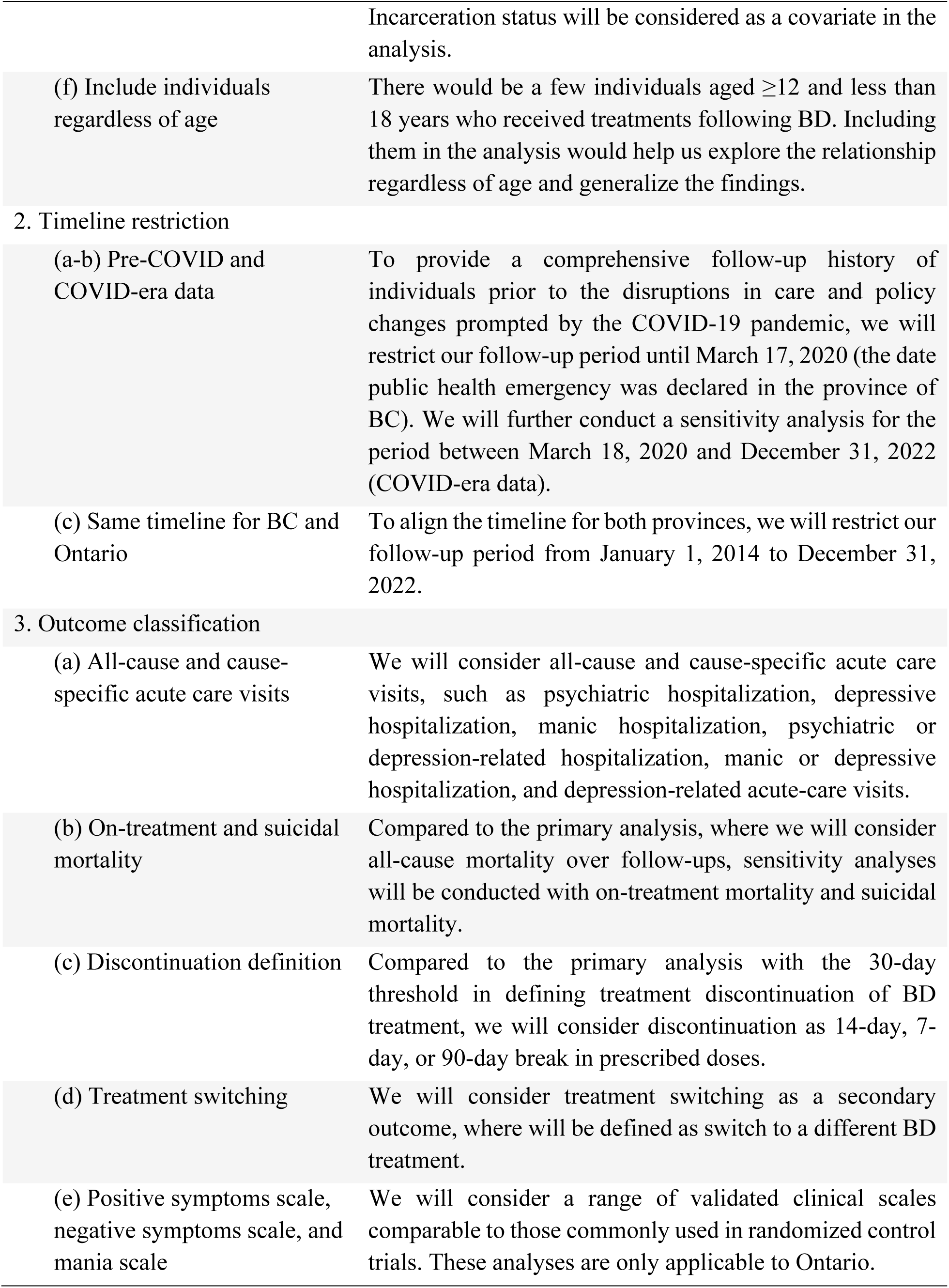
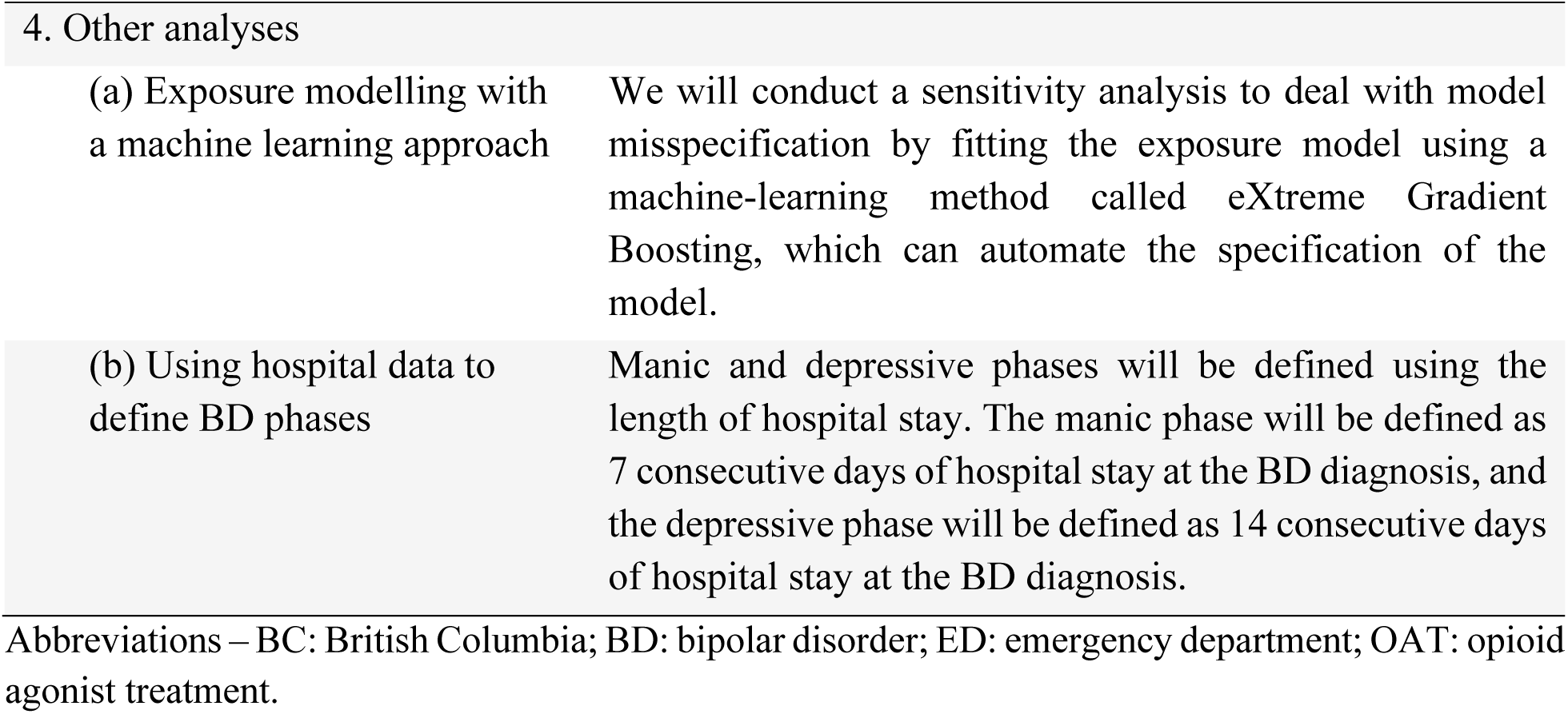
Proposed subgroup and sensitivity analyses in determining the comparative effectiveness of treatment options for bipolar disorder among people with opioid use disorder.

| Sensitivity and subgroup analyses | Rationale |
| --- | --- |
| 1. Sample restriction |  |
| (a) Alternative washout period restrictions | To accommodate the impact of the 30-day washout period in defining prevalent new users, we will consider 60- and 90-day washout periods. |
| (b) Exclude individuals not on OAT | Individuals not on OAT may be systematically different from those on OAT. Our subgroup analysis will exclude those who are not on OAT at BD treatment initiation. Another subgroup analysis will be conducted only including those who are not on OAT. |
| (c) Exclude individuals getting care from community health centres | Individuals receiving care from community health centres are often clinically unstable and have multiple comorbidities. Our sensitivity analysis will exclude these individuals getting care from community health centres at time zero. |
| (d) Exclude individuals with schizophrenia | Individuals diagnosed with schizophrenia are likely to experience more severe symptoms than those without. Our primary analysis excludes individuals who are diagnosed with schizophrenia within one year before time zero. Sensitivity analyses will be conducted by excluding individuals with schizophrenia within five years before time zero as well as excluding lifetime schizophrenia. |
| (e) Include individuals regardless of incarceration status | To align the analyses for both provinces, we will include individuals regardless of incarceration status in BC. |
|  | Incarceration status will be considered as a covariate in the analysis. |
| (f) Include individuals regardless of age | There would be a few individuals aged $\geq 12$ and less than 18 years who received treatments following BD. Including them in the analysis would help us explore the relationship regardless of age and generalize the findings. |
| 2. Timeline restriction |  |
| (a-b) Pre-COVID and COVID-era data | To provide a comprehensive follow-up history of individuals prior to the disruptions in care and policy changes prompted by the COVID-19 pandemic, we will restrict our follow-up period until March 17, 2020 (the date public health emergency was declared in the province of BC). We will further conduct a sensitivity analysis for the period between March 18, 2020 and December 31, 2022 (COVID-era data). |
| (c) Same timeline for BC and Ontario | To align the timeline for both provinces, we will restrict our follow-up period from January 1, 2014 to December 31, 2022. |
| 3. Outcome classification |  |
| (a) All-cause and cause-specific acute care visits | We will consider all-cause and cause-specific acute care visits, such as psychiatric hospitalization, depressive hospitalization, manic hospitalization, psychiatric or depression-related hospitalization, manic or depressive hospitalization, and depression-related acute-care visits. |
| (b) On-treatment and suicidal mortality | Compared to the primary analysis, where we will consider all-cause mortality over follow-ups, sensitivity analyses will be conducted with on-treatment mortality and suicidal mortality. |
| (c) Discontinuation definition | Compared to the primary analysis with the 30-day threshold in defining treatment discontinuation of BD treatment, we will consider discontinuation as 14-day, 7-day, or 90-day break in prescribed doses. |
| (d) Treatment switching | We will consider treatment switching as a secondary outcome, where will be defined as switch to a different BD treatment. |
| (e) Positive symptoms scale, negative symptoms scale, and mania scale | We will consider a range of validated clinical scales comparable to those commonly used in randomized control trials. These analyses are only applicable to Ontario. |
| 4. Other analyses |  |
| (a) Exposure modelling with a machine learning approach | We will conduct a sensitivity analysis to deal with model misspecification by fitting the exposure model using a machine-learning method called eXtreme Gradient Boosting, which can automate the specification of the model. |
| (b) Using hospital data to define BD phases | Manic and depressive phases will be defined using the length of hospital stay. The manic phase will be defined as 7 consecutive days of hospital stay at the BD diagnosis, and the depressive phase will be defined as 14 consecutive days of hospital stay at the BD diagnosis. |

Abbreviations – BC: British Columbia; BD: bipolar disorder; ED: emergency department; OAT: opioid agonist treatment.

### Outcomes

The primary outcomes of interest are the time to any psychiatric acute-care visits, discontinuation of BD treatment and all-cause mortality. The secondary outcomes are all-cause and cause-specific acute-care visits, all-cause and cause-specific hospitalizations, all-cause and cause-specific emergency department visits, BD treatment switching, on-treatment mortality, suicide mortality, positive symptoms scale, negative symptoms scale, and mania scale. Acute care visits will be observed via hospitalizations or emergency department visits and mortality information will be observed via linkage to vital statistics records. Treatment discontinuation will be observed via drug dispensation records, which will be defined as a break in prescribed doses lasting thirty or more days. We chose the 30-day threshold since it is the most common definition for discontinuation of BD treatment based on medication fills.^102–105^ Sensitivity analyses will be conducted by defining discontinuation as a 7-day, 14-day, or 90-day break in prescribed doses, as seen in previous studies.^102,105^ Treatment switching will be defined as switch to a different BD treatment compared to the observed BD treatment at time zero. Positive symptoms scale, negative symptoms scale, and mania scale will be defined using the RAI-MH version 2.0.^106,107^

### Analysis plan

Our aim is to determine the effectiveness of pharmacological treatment options for BD in people with OUD in reducing acute mental health care visits, improving treatment retention, and delaying mortality. We propose to conduct initiator (intention-to-treat conditional on medication dispensation) and per-protocol analyses in parallel, in both BC and Ontario. An ‘initiator analysis’ that allows flexible dosing schedules set by prescribing physicians will focus on an individual’s outcome at death or at the end of follow-up, adjusting for investigator-specified measured confounders. We will also implement high-dimensional propensity score (hdPS) and instrumental variable (IV) analyses to deal with bias due to unmeasured confounding.^108,109^ However, in the presence of frequent treatment switching and suboptimal dosing, the initiator treatment effect might be less meaningful for clinical decision making. A longitudinal per-protocol (PP) analysis, in which clients will be censored once they explicitly deviate from the study protocol for each medication, will be conducted to estimate the comparative effectiveness of each medication regimen after adjusting for treatment switching. An additional PP analysis will be conducted for the first and second sets of treatment strategies to estimate the comparative effectiveness when offered at the recommended dose per clinical guidelines. Details are below.

#### Initiator analysis approach

##### Inverse probability weighting with investigator-specified covariates

We will apply the inverse probability weighting (IPW) approach using investigator-specified covariates to control for potential selection bias in each medication regimen. There are four steps in the IPW approach. First, we will fit the multinomial logistic regression with the investigator-specified covariates to estimate the propensity scores. Second, we will calculate the stabilized inverse probability weights to prevent extreme weights.^110^ If we observe extreme weights, we will truncate them at a smaller level such as at the 99th percentile to avoid undue influence of outliers. Third, we will assess the distribution of the covariates by treatment strategy in the IP-weighted pseudo-population using standardized mean differences (SMDs). An SMD of less than 0.1 will be considered as good covariate balancing.^111^ Fourth, we will fit the Cox proportional hazards (Cox-PH) model to the inverse-probability-weighted population. Covariates with SMD ≥ 0.1 in step 3 will be included in the model. We will report the adjusted hazard ratio (aHR) with the 95% compatibility/confidence interval (95% CI). White’s robust sandwich estimator will be used to calculate the SE and 95% CI.^112^ We will also estimate the adjusted risk difference (aRD) and cumulative incidence curves, with 200 bootstrap samples to obtain the 95% CI.^113^

##### hdPS analysis with high-dimensional proxy data

A systematic review reported that baseline disease severity and treatment resistance are measured and accounted for poorly in RCTs.^114^ With the wealth of high-dimensional health administrative data, we will conduct the hdPS analysis to minimize residual confounding bias.^108^ There are seven major steps in the hdPS analysis.^108,115^ First, proxy information will be obtained from three linked data sources with a one-year covariate assessment period prior to the time zero: (i) physician visits: 3-digit ICD-9 diagnostic codes, (ii) hospitalizations: 3-digit ICD-9/10 diagnosis codes and procedure codes, and (iii) prescriptions dispensed: drug identification numbers. Second, ICD-9/10 codes or drug identification numbers that are part of the investigator-specified covariates or treatment strategies will be excluded to avoid double counting. Third, recurrence of proxies/codes will be converted into binary variables (‘empirical covariates’). Fourth, Cox-PH with LASSO regularization will be used to rank the empirical covariates (**Appendix-A**). Fifth, the top 200 covariates prioritized by log-HR will be selected. Sixth, multinomial logistic regression will be fitted with the investigator-specified and 200 empirical covariates. The stabilized weight will be calculated. We will truncate the weights at the 99th percentile if needed. We will check the balance of the investigator-specified covariates in terms of SMD. Seventh, the Cox-PH model on the IP-weighted population will be fitted, and robust sandwich estimator will be used to calculate the SE.

##### IV analysis

We will conduct an IV analysis using the two-stage residual inclusion (2SRI) approach to address potential unmeasured confounding. In the first stage, we will model the exposure using a multinomial logistic regression that includes the investigator-specified covariates and the IV. Consistent with the approach by Homayra et al.,^116^ we will evaluate several prescriber- and region-level prescribing preference measures as potential IVs. Specifically, we will consider (a) prescriber-level preference, defined as the proportion of patients with concurrent BD and OUD who received the target medication from that prescriber in the prior 12 months; and (b) health-authority (HA) or facility-level preference, defined analogously using aggregated prescribing patterns within the HA or facility. Each candidate IV will be assessed against the four standard IV assumptions, including relevance, exclusion restriction, independence, and monotonicity. We will perform sensitivity analyses using alternative IV definitions (e.g., categorical vs continuous formulations, prescriber- vs region-level) to assess the robustness of estimates. Since we are dealing with a categorical exposure, we will compute generalized residuals as proposed by Gourieroux et al.^117^ In the second stage, we will fit the Cox-PH model an outcome model that includes the exposure, investigator-specified covariates, and the generalized residuals from the first stage. We will use White’s robust sandwich estimator to calculate the SE and 95% CI.

#### Per-protocol approach

Individuals could switch to a different medication if there is a poor response after two months on a recommended dose.^12^ Also, the recommended doses could be different at different stage of the treatment. To compare the one-to-one BD treatment options (i.e., the first and second sets of treatment strategies), we will conduct two PP analyses to deal with (i) treatment switching and (ii) treatment options at recommended doses. For the third and fourth sets of treatment strategies, we plan to conduct a PP analysis to deal with treatment switching as guidelines on recommended doses for combinations of treatments are unavailable.

##### Per-protocol analysis on treatment switching

We will implement the marginal structural modelling (MSM) approach to deal with treatment switching. We will use pooled multinomial logistic regression with inverse probability of censoring weights to generate PP estimates controlling for time-varying confounding. In addition to treatment weights, we will calculate two sets of censoring weights using pooled logistic regression, one for artificial censoring due to switching (where individuals will be censored when they switch to a different treatment strategy) and other one for censoring due to deviation from baseline eligibility criteria (i.e., diagnosed with kidney or liver toxicity, received cancer treatment or palliative care, becoming pregnant, incarcerated, or out-migration from the province). Three sets of weights will be multiplied. Stabilized weights will be calculated and truncated at the 99th percentile if needed. We will use White’s robust sandwich estimator to calculate the 95% CI.

##### Per-protocol analysis on dose adjustment

In the dose adjustment analyses in comparing the first and second sets of treatment strategies, we will compare the treatments at recommended dosage, which will be defined as receipt of at least one dispensation of the following dosages: lithium (≤300 mg/day),^118–120^ divalproex (500-750 mg/day),^121^ lamotrigine (25 mg/day),^122,123^ valproic acid (500-750 mg/day),^121^ risperidone (1-3 mg/day),^124,125^ olanzapine (5-20 mg/day),^126^ aripiprazole (10-30 mg/day),^127–129^ and quetiapine (300-600 mg/day)^130^ (**Appendix Table 5**). The MSM approach will be used. In addition to treatment weights estimated using pooled multinomial logistic regression, we will calculate three sets of censoring weights with pooled logistic regression: artificial censoring to deal with dose adjustment (where individuals will be censored when they deviate from their initial dose), artificial censoring to deal with treatment switching and censoring to deal with deviation from baseline eligibility criteria. Stabilized weights will be calculated, and four sets of weights will be multiplied. We will truncate the weights at the 99th percentile if needed. White’s robust sandwich estimator will be used to calculate the SE and the 95% CI.

#### Covariates

We conducted a systematic literature review of articles published up to 31 May 2025 to identify factors associated with psychiatric acute-care visits, depression-related acute-care visits, and all-cause mortality (**Appendix-B**). The search was restricted to studies on humans, reported in English, and published after 31 December 2000 to ensure the findings were relevant to current treatment options. A total of 94 articles were identified through this search and screened for inclusion. **Table 3** shows the list of covariates. We will supplement this list with additional covariates that are hypothetically relevant for the analyses with people with concurrent OUD and BD,^131^ such as variables controlling for whether taking treatments for OUD, as well as timing variables, including calendar year. However, we will exclude IVs, mediators, and colliders from the analyses (**Appendix Figure 1**). Time-fixed covariates will be defined at time zero. Time-dependent covariates that will be used only for the PP analyses will be defined in a one-year look-back period and updated weekly.

#### Sensitivity and subgroup analyses

We will conduct a range of sensitivity and subgroup analyses to assess the robustness of our findings and heterogeneity in treatment effects across subgroups. Our sensitivity and subgroup analyses will include cohort restriction, timeline restriction, outcome classification, and propensity score model specification^132^ (**Table 4**). To conduct the phase-specific comparison across both provinces, we will conduct a sensitivity analysis by defining manic and depressive phases based on the length of hospital stay. The manic phase will be defined as 7 consecutive days of hospital stay at the BD diagnosis, and the depressive phase will be defined as 14 consecutive days of hospital stay at the BD diagnosis.^133^ Given the limitations on population-level data capture in Ontario, we will otherwise restrict the population in BC to provide a more direct comparison to Ontario. Any deviations from this protocol will be noted in the final report.

### Ethics and dissemination

The linked databases were made available to the research team by British Columbia Ministries of Health and Mental Health and Addiction as part of the response to the provincial opioid overdose public health emergency. The study was classified as a quality improvement initiative. Providence Health Care Research Institute and the Simon Fraser University Office of Research Ethics determined the analysis met criteria for exemption per Article 2.5 of the Tri-Council Policy Statement: Ethical Conduct for Research Involving Humans.^134^

For conducting and reporting research, this study will adhere to international guidelines of the Transparent Reporting of Observational Studies Emulating a Target Trial (The TARGET Statement).^135^ Results will be disseminated to decision-makers, local advocacy groups, and national and international clinical guideline developers. The study findings will also be presented at international conferences and published in peer-reviewed journals. Overall, this study will generate robust evidence regarding the effectiveness of pharmacological treatment options for BD among people with OUD as seen in real-world clinical practice, in the interest of improving retention in this essential^136^ and life-saving^137,138^ BD medication.

### Patient and public involvement

No patients were directly involved in designing this study. The findings will be shared with local advocacy groups following completion of the analysis.

## Data Availability

Access to data provided by the Data Stewards is subject to approval but can be requested for research projects through the Data Stewards or their designated service providers. The following data sets were used in this study: PharmaNet, Client Roster, Medical Services Plan, Discharge Abstract Database, National Ambulatory Care Reporting System Database, Provincial Corrections, Perinatal Database, Social Development and Poverty Reduction Database, and Vital Statistics. All inferences, opinions, and conclusions drawn in this publication are those of the author(s), and do not reflect the opinions or policies of the Data Steward(s). This Data was provisioned under ISP 17-162.

https://www2.gov.bc.ca/gov/content/health/conducting-health-research-evaluation/data-access-health-data-central

## Contributors

BN and DM conceptualized and secured funding for the study. MBH wrote the first draft of the article and led the review and methodological development. All authors provided critical revisions to the manuscript and approved the final draft. BN is responsible for the overall content as the guarantor.

## Role of the Funding source

The funding source was independent of the design of this study and did not have any role during its execution, analyses, interpretation of the data, writing or decision to submit results. The authors had full access to the results in the study and took responsibility for the integrity of the data and accuracy of the analysis.

## Conflict of interests

None declared.

## Notes

### Competing Interest Statement

The authors have declared no competing interest.

### Funding Statement

This study was funded by the Canadian Institutes of Health Research (Grant #195860).

