## Appendix for "Comparative effectiveness of preferred pharmacological treatment options for bipolar disorder among people with opioid use disorder in British Columbia and Ontario, Canada: protocol for parallel population-based target trial emulations"

Hossain et al.

### Appendix-A: Top 200 empirical covariates using Cox-LASSO

To identify the top 200 empirical covariates using Cox proportional hazard (Cox-PH) model with LASSO regularization (Cox-LASSO), we will follow the steps:

- Fit the Cox-LASSO shrinkage as the outcome model with all empirical covariates. The lambda hyperparameter will be selected using 5-fold cross-validation. We will select the lambda value that results in the minimum prediction error.
- Extract the log hazard ratio (HR) for each empirical covariate.
- Rank empirical covariates based on the absolute log-HR.
- Select the top 200 empirical covariates.

### Appendix-B: Review of factors associated with psychiatric acute-care visits, depression-related acute-care visits and all-cause mortality for people with bipolar disorder

#### Search terms including mesh

*Health service outcomes:* ("Bipolar Disorder" OR bipolar OR "manic depress*) AND ("Risk Factors" OR predictor* OR determinant* OR correlate* OR association* OR "hazard ratio" OR "odds ratio" OR "risk ratio" ) AND ("Emergency Service, Psychiatric" OR "Emergency Service, Hospital" OR "Hospitalization" OR "Patient Readmission" OR "emergency department" OR "psychiatric emergency" OR "emergency psychiatry" OR "mental health crisis" OR "crisis unit" OR "crisis stabilization" OR "crisis intervention" OR hospitaliz* OR readmit* OR "unplanned visit" OR "unplanned visits" OR "acute care" )

*Mortality outcome:* ("Bipolar Disorder" OR bipolar OR "manic depress*") AND ("Risk Factors" OR predictor* OR determinant* OR correlate* OR association* OR "hazard ratio" OR "odds ratio" OR "risk ratio") AND ("Mortality" OR "Case Fatality Rate" OR "Suicide" OR mortality OR death* OR fatal*)

#### Population

Adults diagnosed with bipolar disorder (BD I/II) by clinical record or validated criteria (DSM/ICD).

#### Exposure(s)

Factor associated with the outcomes (e.g., demographics, comorbidities incl. SUD, treatment patterns, healthcare access, social determinants).

#### Comparator

Within-study comparators: presence vs absence of the factor; different categories/levels

#### Outcomes

- *Psychiatric-related acute-care visits* (ED/psychiatric emergency service, crisis unit, unplanned admissions/readmissions)
- *Depression-related acute-care visits* (primary reason/diagnosis is depressive episode)
- *All-cause mortality* (inpatient, post-discharge, or population-level)

#### Eligibility criteria

- *Inclusion:* Human studies; BD explicitly defined; any country; publication year unrestricted; English or French
- *Exclusion:* Case reports/series (<10); editorials; studies with mixed severe mental illness where BD results aren’t separable; intervention trials reporting only treatment effects without factor–outcome associations; studies focused solely on non-psychiatric acute care without psychiatric or depression-related endpoints.

**Results**

| Covariate | Association | Quality of evidence† (source) |
|---|---|---|
| Individual-related characteristics | | |
| Age (18+) | +Psychiatric-related, All-cause mortality[1,2] | Level II |
| Gender (male) | +All-cause mortality[3] | Level II |
| Living in a rural area | + Depression-related[4] | Level II |
| Ethnicity (minority groups) | +All-cause mortality[3] | Level II |
| Homelessness | +All-cause mortality[2] | Level II |
| Socioeconomic status (high vs low) | +All-cause mortality[3] | Level II |
| Concurrent conditions | | |
| SUD history | +Psychiatric-related[1,5] | Level II |
| Type of substance use | +All-cause mortality[5,6] | Level II |
| Lung function decline / respiratory impairment | +All-cause mortality[7] | Level II |
| Healthcare/Medication history | | |
| Lithium treatment | -All cause (suicide deaths)[8] | Level I |
| Antipsychotic selection (lurasidone vs other atypicals) | -Psychiatric[9] | Level II |
| Healthcare utilization history (prior ED/psychiatric admissions) | +Psychiatric[1,10] | Level II |
| Psychoeducation program participation | +Psychiatric[2,4] | Level II |

*Quality of evidence ratings: level I: systematic reviews, meta-analyses and randomized controlled trials; level II: cohort studies, case–control studies, case studies; level III: case reports, ideas, editorials, opinions (source: Cochrane review library).*

*+ = positive association; – = protective association*

**Summary**

A total of 94 records were identified through database searching (PubMed). After removing duplicates (n = 7), 87 titles and abstracts were screened. 64 were excluded at the title/abstract stage for not meeting inclusion criteria (e.g., non-BD populations, non-relevant outcomes, or ineligible designs). Remaining 23 full-text articles were assessed for eligibility. 13 were excluded due to mixed serious mental illness (SMI) populations with non-separable BD results (n = 6), absence of acute-care or mortality outcomes (n = 4), or ineligible study designs such as case series and qualitative reports (n = 3).

10 studies met inclusion criteria and were included in Table 1. These included 2 randomized controlled trials (Level I evidence) and 8 observational cohort studies (Level II evidence), representing populations across North America, Europe, and Asia.

Consistent predictors of psychiatric acute-care use emerged. Substance use disorder (SUD) history was a major risk factor for increased hospitalization or readmission.[1,5] Prior psychiatric hospitalizations or emergency department (ED) visits similarly predicted early-unplanned readmission.[1] In contrast, participation in psychoeducation programs and treatment with lurasidone were protective, each associated with reduced risk of psychiatric admission or ED presentation.[4,9] The use of electroconvulsive therapy (ECT) during manic episodes was also linked with longer time to readmission, indicating a potential stabilizing effect on acute-care trajectories.[10]

Mortality-related outcomes were primarily informed by population-based registry and clinical trial data. Ethnic minority status and socioeconomic deprivation were associated with significantly reduced life expectancy and higher all-cause mortality among individuals with serious mental illness, including those with BD.[3] Homelessness was another strong predictor of premature death within the Housing First randomized controlled trial.[2] Physical health factors, notably lung-function decline and respiratory impairment, were also associated with increased mortality risk.[7] While lithium treatment was not directly evaluated for acute-care outcomes, it demonstrated a protective signal for repeat suicide-related mortality in a mixed veteran cohort with major depression or BD.[8]

Overall, evidence indicates that psychosocial vulnerability (homelessness and SUD), prior service utilization, and untreated comorbidity are major drivers of acute-care use and mortality in BD. Conversely, structured psychoeducation reduces acute-care burden. The collective findings underscore the need for integrated approaches addressing both clinical and social determinants of health in this population.

This review identified multiple studies examining predictors of psychiatric hospitalization, readmission, and mortality among individuals with BD. There was no study specifically analyzing depression-related acute-care visits as a separate outcome. Existing literature combines mood-episode–related admissions or ED presentations into a single “psychiatric” category, without distinguishing depressive, manic, or mixed episodes. This shows an important knowledge gap, as depressive episodes are both the most prevalent and the most disabling phase of BD. The absence of depression-specific acute-care data limits understanding of episode-specific drivers of hospital utilization. Future research should aim to disaggregate acute-care outcomes by mood polarity to clarify whether risk and protective factors differ between manic- and depression-related presentations, thereby informing more targeted prevention and service-planning strategies for individuals with BD.

### Appendix Figures

**Appendix Figure 1:** Diagram showing the relationship between pharmacological treatment options for bipolar disorder among people with opioid use disorder (*A*) and outcomes (*Y*).

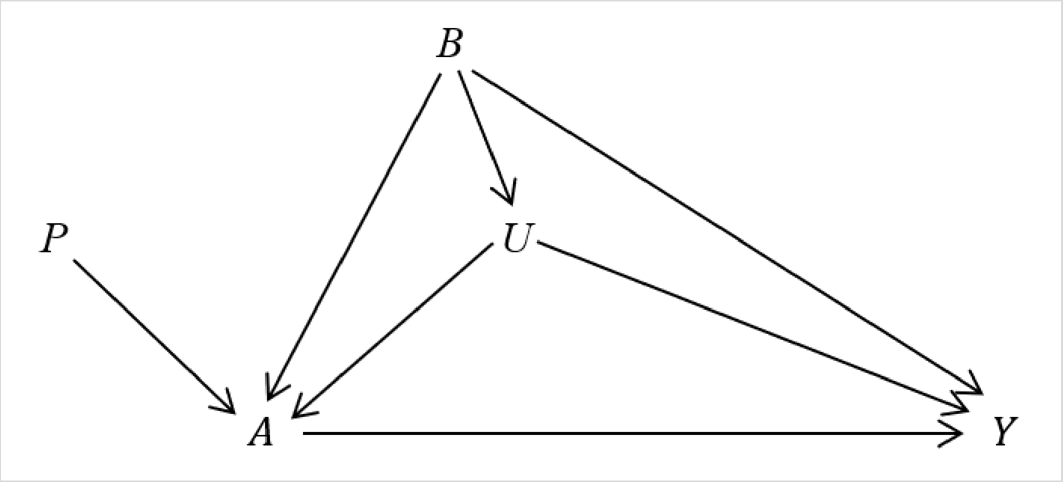

Legend – *A* represents pharmacological treatment options for bipolar disorder (BD), and *Y* represents the study outcomes (acute-care visits, discontinuation, and all-cause mortality). *P* represents instrumental variables such as prescribers' preference to prescribe BD treatments, and *U* represents unmeasured covariates such as severity of BD and BD treatment-resistance. Baseline/time-fixed confounders *B* include age, sex, place of residence, unstable housing, receipt of income assistance, incarceration status, indication of non-opioid substance use disorder, alcohol use disorder, tobacco use disorder, and respiratory impairment, Charlson comorbidity index, chronic disease score, calendar year, hepatitis C, chronic pain, antipsychotic for BD, history of healthcare utilization, time since first indication of BD, long-acting injectable, receipt of other mental health medications, major depressive disorder, whether received sedative medication, opioid dose, cumulative exposure to opioid agonist treatment, number of drug-related emergency department visits, and care from community health centres.

### Appendix Tables

**Appendix Table 1:** Guideline comparison of first and second-line pharmacological treatment options for bipolar disorder.

| Guideline | Recommendation on treatment options following BD | Recommendation for people with OUD | Supporting references |
|---|---|---|---|
| ISBD (2009)[11] | First-line (acute): Lithium, Quetiapine, Lamotrigine, Adjunctive antidepressants.<br>First-line (maintenance): Lithium, Divalproex, Lamotrigine, Quetiapine, Olanzapine.<br>Second-line (acute): Adjunctive antidepressants, Electroconvulsive therapy.<br>Second-line (maintenance): Carbamazepine, Adjunctive atypical antipsychotics, Combination such as Lithium + Divalproex. | None. | Baldessarini et al. (2002)[12], Goodwin et al. (2003)[13], Cipriani et al. (2005)[14], Carney and Goodwin (2005)[15], Scherk et al. (2007)[16], Jarema et al. (2007)[17], Azorin and Findling (2007)[18], Vitiello et al. (1998)[19], Oudit et al. (2007)[20], American Psychiatric Association (2002)[21], NICE (2006)[22], CANMAT (2005)[23]. |
| American Psychiatric Association, US (2002)[21] | First-line for bipolar I (acute): Lithium, Valproate, Lithium + antipsychotic, Valproate + antipsychotic; Lithium and Lamotrigine for depressive episodes.<br>First-line for bipolar I (maintenance): Lithium, Valproate,<br>First-line for bipolar II (acute): Lithium, Lamotrigine.<br>First-line for bipolar II (maintenance): Lithium, Valproate. | No specific treatment recommendations, but lithium and lamotrigine could be preferred due to mood-stabilizing effects. | Tondo and Baldessarini (2000)[24], Ahrenset al. (1995)[25], Tondo et al. (1997)[26], Goodwin et al. (1969)[27], Cundall et al. (1972)[28], Coppen et al. (1973)[29], Baastrup et al. (1970)[30], Bowden et al. (2000)[31], Lambert and Venaud (1992)[32], Calabrese et al. (2001)[33], Greil et al. (1997)[34], Esparon et al. (1986)[35]. |

| | | | |
|---|---|---|---|
| | Second-line for bipolar I (acute): Carbamazepine or Oxcarbazepine, Quetiapine or Ziprasidone, Clozapine, Electroconvulsive.<br><br>Second-line for bipolar I (maintenance): Lamotrigine, Carbamazepine, or Oxcarbazepine.<br><br>Second-line for bipolar II (acute): Antidepressants, Lithium + antidepressant.<br><br>Second-line for bipolar II (maintenance): Lamotrigine, Carbamazepine, or Oxcarbazepine. | | |
| CANMAT and ISBD, Canada (2018)[36] | First-line for bipolar I (acute): Quetiapine, Lithium, Lamotrigine and Lurasidone are all recommended as monotherapy. Lurasidone and Lamotrigine are also recommended as adjunctive treatments (adj). Quetiapine + Lithium in patients who experience depression.<br><br>First-line for bipolar I (maintenance): Lithium, Quetiapine, Divalproex, Lamotrigine, Asenapine, Quetiapine + Lithium/Divalproex (Li/DVP), Aripiprazole, Aripiprazole + Li/DVP.<br><br>First-line for bipolar II: Quetiapine.<br><br>Second-line for bipolar I (acute): Monotherapy with divalproex, and adjunctive use of antidepressant therapy (selective serotonin reuptake inhibitors or bupropion) with Li/DVP or an atypical antipsychotic as an add-on treatment. | Lithium and lamotrigine may be effective. | Severus et al. (2014)[37], Miura et al. (2014)[38], Weisler et al. (2011)[39], Cipriani et al. (2013)[40], Bowden et al. (2000)[31], Calabrese et al. (2006)[41], Szegedi et al. (2018)[42], Keck et al. (2006)[43], Calabrese et al. (2017)[44], Suppes et al. (2009)[45], Vieta et al. (2008)[46], Tohen et al. (2006)[47], Vieta et al. (2012)[48], Macfadden et al. (2009)[49], Post et al. (2002)[50], Berwaerts et al. (2012)[51], Bowden et al. (2010)[52], Calabrese et al. (2017)[53], Geller et al. (1998)[54] |

| | | | |
|---|---|---|---|
| | Second-line for bipolar I (maintenance): Olanzapine, Risperidone, Carbamazepine, Paliperidone, Lurasidone + Li/DVP, Ziprasidone + Li/DVP.<br><br>Second-line for bipolar II: Lithium, Lamotrigine, Bupropion (adj), Electroconvulsive, Sertraline, Venlafaxine. | | |
| CANMAT and ISBD, Canada (2024)[55] | First-line for bipolar I (acute): Quetiapine, lurasidone + Li/DVP, Lithium, Lamotrigine, Cariprazine, Lurasidone (adj).<br><br>First-line for bipolar I (maintenance): Lithium, Quetiapine, Divalproex, Lamotrigine, Asenapine, Quetiapine + Li/DVP, Aripiprazole + Li/DVP, Aripiprazole.<br><br>First-line for bipolar II (acute): Quetiapine.<br><br>First-line for bipolar II (maintenance): Quetiapine, Lithium, Lamotrigine.<br><br>Second-line for bipolar I (acute): Divalproex, Selective serotonergic reuptake inhibitor/Bupropion (adj), Electroconvulsive therapy, Olanzapine-fluoxetine, Lumateperone.<br><br>Second-line for bipolar I (maintenance): Olanzapine, Risperidone, Risperidone (adj), Carbamazepine, Paliperidone, Lurasidone + Li/DVP, Ziprasidone + Li/DVP. | Lithium may be effective. | CANMAT and ISBD, Canada (2018)[36], Geller et al. (1998)[54], Kishi et al. (2021)[56], Calabrese et al. (2021)[57]. |

| | | | |
|---|---|---|---|
| | Second-line for bipolar II (acute): Lithium, Lamotrigine, Bupropion (adj), Electroconvulsive therapy, Sertalinea, Venlafaxinea, Lumateperone.<br><br>Second-line for bipolar II (maintenance): Venlafaxine. | | |
| CAMH, Canada (2013)[58] | Mood stabilizers: Lithium, Divalproex, Valproic acid or Valproate, Carbamazepine, Lamotrigine.<br><br>Antidepressant medications: Selective serotonin reuptake inhibitors, Serotonin and norepinephrine reuptake inhibitors, Norepinephrine and Dopamine reuptake inhibitors, Noradrenergic and specific serotonergic antidepressants, Cyclics, Monoamine oxidase inhibitors, Anti-anxiety medications.<br><br>Antipsychotic medications: Risperidone, Quetiapine, Olanzapine, Ziprasidone, Paliperidone, Aripiprazole, Clozapine, Chlorpromazine, Flupenthixol, Fluphenazine, Haloperidol, Loxapine, Perphenazine, Pimozide, Trifluoperazine, Thiothixene, And Zuclopenthixol, Electroconvulsive therapy. | None. | None. |
| Early Psychosis Guide, BC, Canada (2002)[59] | Antipsychotic and either Lithium or Valproate; Benzodiazepines if severe behavioural disturbance is present; second mood stabilizer (e.g., Olanzapine and Risperidone) or switching antipsychotics if poor response after three weeks.[60–67] | Clozapine can be effective in reducing both psychotic symptoms and substance abuse.[68–71] | APA guideline[60], Vieta et al. (2001)[61], Tohen et al. (1996)[62], McElroy et al. (1998)[63], Tohen et al. (2000)[64], Sachs et al. (2000)[65], Bauer et al. (1999)[66], Vieta et al. (2001)[67], Buckley et al. (1994)[68], McElroy et al. (1995)[69], Drake et al (2000)[70], Zimmet et al. (2000)[71] |

| | | | |
|---|---|---|---|
| Family physician guide, BC, Canada (2007)[72] | Mood stabilizer: Lithium, Valproate, Carbamazepine, Lamotrigine, Gabapentin.<br><br>Antipsychotics: Risperidone, Olanzapine, Quetiapine. | No specific treatment recommendations. | CANMAT guideline[73], Goodwin (2003)[74], APA guideline[60] |
| Reproductive Mental Health Program & Perinatal Services, BC, Canada (2014)[75] | Lithium, Carbamazepine, Gabapentin, Lamotrigine, Topiramate, and Valproic acid/Divalproex | No specific treatment recommendations. | Cohen (2007)[76], Viguera (2000)[77], Freeman et al. (2002)[78], Sylvia et al. (2008)[79], Miklowitz (2008)[80], Benyon et al. (2008)[81], Scott et al. (2007)[82] |
| National Institute for Health and Care Excellence, UK (2014)[83] | Lithium, Valproate, Lamotrigine, and Antipsychotic medication.<br><br>Adults in primary care: Do not start valproate; Do not start lithium if lithium is not taken before.<br><br>Adults in secondary care: Offer lithium as a first-line. If lithium is ineffective, consider adding valproate. Offer Haloperidol, Olanzapine, Quetiapine or Risperidone if not taking an antipsychotic or mood stabilizer. | None. | None. |
| National Institute for Health and Care Excellence, UK (2023)[84] | Lithium, Valproate, Lamotrigine, and Antipsychotic medication.<br><br>Adults in primary care: Do not start valproate; Do not start lithium if lithium is not taken before.<br><br>Adults in secondary care: Offer lithium as a first-line. If lithium is ineffective, poorly tolerated, or is not suitable, consider an antipsychotic such as Asenapine, Aripiprazole, Olanzapine, Quetiapine or | None. | National Institute for Health and Care Excellence, UK (2014)[83]. |

| | | | |
|---|---|---|---|
| | Risperidone; If the first antipsychotic is poorly tolerated at any dose or ineffective at the maximum licensed dose, consider an alternative antipsychotic; If an alternative antipsychotic is ineffective, consider a combination of valproate with either an antipsychotic or lithium. | | |
| British Association for Psychopharmacology (2016)[85] | Bipolar I/II (acute): Quetiapine, Olanzapine, Olanzapine + Fluoxetine, Antidepressants, Lurasidone, Lamotrigine as combination.<br><br>Bipolar I/II (maintenance): Lithium, Dopamine antagonists and partial agonists, Valproate, Lamotrigine. | Lithium + Valproate. | National Institute for Health and Care Excellence, UK (2014)[83], Kemp et al. (2009)[86]. |
| National Health Service, UK (2023)[87] | Bipolar I/II (acute): If there is no history of taking lithium or valproate, start with Olanzapine + Fluoxetine, Quetiapine, or Olanzapine. If there is a history of taking lithium or valproate, use optimise mood stabilizer alongside Olanzapine + Fluoxetine, Quetiapine, or Olanzapine. If ineffective, consider Lamotrigine. If it is still Ineffective/ not tolerated consider Aripiprazole, Lurasidone, or Electroconvulsive therapy.<br><br>Bipolar I/II (maintenance): Lithium. if poorly tolerated/unsuitable, replace with Valproate, Olanzapine, or Quetiapine. If lithium is ineffective, add valproate or atypical antipsychotic. | None. | National Institute for Health and Care Excellence, UK (2014)[83], British Association for Psychopharmacology (2016)[85], CANMAT and ISBD, Canada (2018)[36]. |

| | | | |
|---|---|---|---|
| Australian Family Physician, Australia (2013)[88] | Bipolar I/II (acute): Lithium, Valproate, Carbamazepine; First-generation antipsychotics: Chlorpromazine, Haloperidol; Second-generation antipsychotics: Aripiprazole, Asenapine, Olanzapine, Quetiapine, Risperidone, Ziprasidone.<br><br>Bipolar I/II (maintenance): Lithium, Carbamazepine, Lamotrigine; Second generation antipsychotics: Aripiprazole, Asenapine, Olanzapine, Quetiapine, Risperidone. | None. | Geddes et al. (2004)[89], Yatham et al. (2013)[90], Tohen et al. (2006)[47], Vieta et al. (2008)[46], Bowden et al. (2003)[91], Calabrese et al. (2003)[92], Goodwin et al. (2004)[93], Geddes et al. (2009)[94]. |
| Best Practice Advocacy Centre, New Zealand (2014)[95] | Lithium, Valproate, Carbamazepine, Lamotrigine, Atypical antipsychotics such as Olanzapine, Quetiapine, Risperidone, Aripiprazole or Ziprasidone | None. | Isojarvi et al. (2000)[96], Actavis (2013)[97], ISBD (2009)[11]. |

Abbreviations – APA: American Psychiatric Association; BC: British Columbia; BD: Bipolar Disorder; CAMH: Centre for Addiction and Mental Health; CANMAT: Canadian Network for Mood and Anxiety Treatments; ISBD: International Society for Bipolar Disorders; OUD: Opioid use disorder; Li/DVP: Lithium/Divalproex.

**Appendix Table 2:** Clinical trials comparing pharmacological treatment options for bipolar disorder.

| Author(s) | Population | Treatment |
|---|---|---|
| Nierenberg et al. (2014)[98] | Outpatients with BD. People with comorbid substance abuse were included. | Quetiapine versus Lithium, each with adjunctive personalized treatments. |
| McIntyre et al. (2019)[99] | Veterans with BD or depression who had survived a recent suicide-related event. People with substance abuse and/or dependence were excluded. | Extended-release lithium carbonate beginning at 600 mg/d versus Placebo augmentation of usual care. |
| Preskorn et al. (2022)[100] | Adults with BD I or II. | Sublingual dexmedetomidine 180µg, 120µg, or placebo. |
| Mirzazadeh et al. (2025)[101] | People diagnosed with BD I who were experiencing acute mania. People with substance use disorder were excluded. | Levetiracetam with olanzapine versus Placebo with olanzapine. |
| Sahraian, et al. (2018)[102] | People with BD who had obsessive and compulsive symptoms. People with substance use disorder were excluded. | Aripiprazole (lithium 725 mg, clonazepam 0.4 mg and aripiprazole 16.32 mg) versus No aripiprazole (lithium 700 mg, clonazepam 0.5 mg and placebo). |
| Habibi et al. (2017)[103] | Children and adolescents aged 10-18 years who were hospitalized for a BD manic or mixed episode. Children and adolescents with substance use disorder were excluded. | Lithium and quetiapine (400-600 mg per day) versus Lithium and risperidone (0.5-6 mg per day). |
| Oquendo et al. (2011)[104] | Patients with BD and past suicide attempts. People with substance use disorder were included. | Lithium 0.6–1.0 mEq/dl versus Valproate 45–125 µg/ml. Depressed BD patients were randomly assigned to receive either lithium plus paroxetine (up to 60 mg/day if needed) or valproate plus paroxetine. |

| | | |
|---|---|---|
| Sahraian et al. (2014)[105] | People with BD, manic phase type-I, and obsessive-compulsive disorder symptoms. People with substance use disorder were excluded. | Topiramate (lithium, olanzapine, clonazepam, and topiramate) versus Placebo (lithium, olanzapine, clonazepam, and placebo) |
| Talaei et al. (2022)[106] | People with BD in the acute manic phase. People with substance abuse were excluded. | Risperidone and oxcarbazepine versus Risperidone and sodium valproate. |
| Juruena et al. (2009)[107] | People with BD while on lithium maintenance treatment | Lithium and carbamazepine versus Lithium and oxcarbazepine |
| Vieta et al. (2008)[108] | People with BD. People with drug abuse were excluded. | Lithium and oxcarbazepine vs. versus Lithium and placebo. |
| Shafti (2018)[109] | Male in-patients with BD I disorder who presented with relapse or new emergence of an episode of acute mania. People with substance abuse were excluded. | Aripiprazole (5 mg uncoated tablets) versus Lithium carbonate (300 mg uncoated tablets). |
| Shahrbabaki et al. (2020)[110] | People with BD I. People with drug abuse were excluded. | Probiotic (lithium oxide, sodium valproate, risperidone, probiotic capsule) versus Placebo (lithium oxide, sodium valproate, risperidone, placebo). |
| Grunebaum et al. (2017)[111] | People with BD and depression. People with substance use were excluded. | Ketamine (0.5 mg/kg) versus Midazolam (0.02 mg/kg). |
| Khalili et al. (2019)[112] | People with BD I with manic or depressive episodes and psychotic feature and with opioid dependency comorbidity. | Buprenorphine (sodium valproate, risperidone, and buprenorphine) versus Placebo (sodium valproate, risperidone and placebo). |
| Shi et al. (2004)[113] | Adults with BD I, most recent episode depressed. People with substance dependence were excluded. | Olanzapine, Olanzapine-fluoxetine combination, versus Placebo |

| | | |
|---|---|---|
| Vieta et al. (2006)[114] | People with BD I and II. | Gabapentin or placebo added to the current treatment (lithium, valproate, carbamazepine, or any combination but not antipsychotics or antidepressants). |
| Mahmoudi-Gharaei et al. (2012)[115] | Children and adolescents with BD I in the manic or mixed phase. Children and adolescents with substance abuse were excluded. | Topiramate (lithium, risperidone, and topiramate) versus valproate sodium (lithium, risperidone, and valproate sodium). |
| Hegerl et al. (2018)[116] | Popel with BD in acute mania phase and suffering from bipolar affective disorders. People with drug dependency or abuse were excluded. | Methylphenidate (20-40mg/day) versus Placebo. |

BD: bipolar disorder; RCT: Randomized controlled trial.

**Appendix Table 3:** Case definition of opioid use disorder and bipolar disorder in the British Columbia and Ontario cohorts.

| Condition | British Columbia | Ontario |
|---|---|---|
| Opioid use disorder [1] | • ICD-9 from DAD, MSP, VS: 304.0, 304.7, 305.5, 965.0, E850.0-2;<br>• ICD-10 from DAD, NACRS, VS: F11, (X42, X44, X62, X64, Y12, or Y14) and (T40.0, T40.1, T40.2, T40.3, T40.4, or T40.6);<br>• Fee item from MSP: 39, 15039, 13013, 13014, 36521;<br>• Indication of heroin/opioid use as risk factor in pregnancy from BCPDR;<br>• DIN from Pharmanet: 999792, 999793, 66999990, 66999991, 66999992, 66999993, 66999997, 66999998, 66999999, 67000000, 67000008, 67000007, 67000005, 67000006, 67000004, 67000003, 67000001, 67000002, 2242962, 2242963, 2242964, 2295695, 2295709, 66999994, 66999995, 66999996, 2408090, 2408104, 2424851, 2424878, 2453908, 2453916, 2468085, 2468093, 22123349, 22123346, 22123347, 22123348, 22123357, 66123367, 2146126, 22123340, 999776. | • ICD-9 from DAD, NACRS: 304;<br>• ICD-10 from DAD, NACRS: F11;<br>• Fee code from OHIP: K682, K683, K684;<br>• DIN from ODB NMS: 2495783, 2495872, 2495880, 2481979, 2241377, 2247694, 2244290, 2394596, 2394618, 9857221, 9857223, 2247694, 2247698, 2247699, 2247700, 2247701, 9850619, 0985177, 9852891, 9857217, 9857218, 9857219, 9857220, 9857223, 2533650, 2533669, 2533677, 2388383, 2388391, 2388405, 2388413, 2247374, 2408090, 2408104, 2424851, 2424878, 2453908, 2453916, 2295695, 2295709, 2468085, 2468093, 2502313, 2517175, 2517183, 2502321, 2502348, 2502356, 2408104, 2184435, 2184443, 2184451, 2242163, 2483084, 2483092;<br>• DSMCODE from OMHRS: 3040, 3055. |
| Bipolar disorder [2] | • ICD-9 from DAD, MSP, NACRS: 296;<br>• ICD-10 from DAD, MSP, NACRS: F30, F31. | • ICD-9 from DAD, NACRS: 296;<br>• ICD-10 from DAD, NACRS: F30, F31;<br>• Diagnosis code from OHIP: 296;<br>• DSMCODE from OMHRS: 296. |
| Bipolar disorder-I [2] | • ICD-9 from DAD, MSP, NACRS: 296.0, 296.1, 296.4-6, 296.8;<br>• ICD-10 from DAD, MSP, NACRS: F31.0-7, F31.9. | • ICD-9 from OMHRS: 296.0, 296.1, 296.4-6, 296.8;<br>• ICD-10 from OMHRS: F31.0-7, F31.9. |

| | | |
|---|---|---|
| Depressive phase [2] | • ICD-9 from DAD, MSP, NACRS: 296.5;<br>• ICD-10 from DAD, MSP, NACRS: F31.3-5. | • ICD-9 from OMHRS: 296.5;<br>ICD-10 from OMHRS: F31.3-5. |
| Manic phase [2] | • ICD-9 from DAD, MSP, NACRS: 296.0, 296.1, 296.4;<br>• ICD-10 from DAD, MSP, NACRS: F31.0-2. | • ICD-9 from OMHRS: 296.0, 296.1, 296.4;<br>• ICD-10 from OMHRS: F31.0-2. |

Abbreviations – BCPDR: BC Perinatal Data Registry; DAD: Discharge Abstract Database (records of hospitalizations); DIN: drug identification number; ICD: International Classification of Diseases; MSP: Medical Service Plan (physician billing records); NACRS: National Ambulatory Care Reporting System (records of emergency visits); OMHRS: Ontario Mental Health Reporting System (standardized data on adult mental health clients); VS: Vital Statistics.

[1] The case definition of opioid use disorder includes 1 opioid agonist treatment drug dispensation, or ≥3 physician claims, or 1 hospital admission, or 1 emergency department visit, or 1 perinatal, or 1 death record.

[2] The case definition includes 1 hospital admission, or 1 emergency department visit, or ≥3 physician claims at least 30 days apart in a 3-year span.

**Appendix Table 4:** The pharmacological treatment options for bipolar disorder in people with opioid use disorder in Canada.

| Drug | Property | DIN/PIN | Brand name & related details [1] | Health Canada approval date [2] |
|---|---|---|---|---|
| Lithium | Mood stabilizer | 00236683 | Carbolith 300 MG CAPSULE | December 1971 |
| | | 02011239 | Carbolith 600 MG CAPSULE | December 1992 |
| | | 02216132 | Pms-Lithium Carbonate - Cap 150mg 150 MG CAPSULE | December 1996 |
| | | 02216140 | Pms-Lithium Carbonate - Cap 300mg 300 MG CAPSULE | December 1996 |
| | | 02242837 | Apo-Lithium Carbonate 150 MG CAPSULE | February 2001 |
| | | 02242838 | Apo-Lithium Carbonate 300 MG CAPSULE | February 2001 |
| | | 02266695 | Lithmax 300 MG TABLET | April 2004 |
| Divalproex | Anticonvulsants (used as mood stabilizer) | 00596418 | Epival 125 MG TABLET | December 1984 |
| | | 00596426 | Epival 250 MG TABLET | December 1984 |
| | | 00596434 | Epival 500 MG TABLET | December 1984 |
| | | 02239698 | Apo-Divalproex 125 MG TABLET | March 1999 |
| | | 02239699 | Apo-Divalproex 250 MG TABLET | March 1999 |
| | | 02239700 | Apo-Divalproex 500 MG TABLET | March 1999 |
| | | 02458926 | Mylan-Divalproex 125 MG TABLET | April 2017 |
| | | 02458934 | Mylan-Divalproex 250 MG TABLET | May 2017 |
| | | 02459019 | Mylan-Divalproex 500 MG TABLET | April 2017 |
| Lamotrigine | Anticonvulsants (used as mood stabilizer) | 02142082 | Lamictal 25 MG TABLET | December 1995 |
| | | 02142104 | Lamictal 100 MG TABLET | December 1995 |
| | | 02142112 | Lamictal 150 MG TABLET | December 1995 |
| | | 02240115 | Lamictal 5 MG TB CHW DSP | June 1999 |
| | | 02245208 | Apo-Lamotrigine 25 MG TABLET | March 2002 |
| | | 02245209 | Apo-Lamotrigine 100 MG TABLET | March 2002 |
| | | 02245210 | Apo-Lamotrigine 150 MG TABLET | March 2002 |
| | | 02246897 | Pms-Lamotrigine 25 MG TABLET | April 2003 |
| | | 02246898 | Pms-Lamotrigine 100 MG TABLET | April 2003 |

| | | | | |
|---|---|---|---|---|
| | | 02246899 | Pms-Lamotrigine 150 MG TABLET | April 2003 |
| | | 02248232 | Teva-Lamotrigine 25 MG TABLET | January 2004 |
| | | 02248233 | Teva-Lamotrigine 100 MG TABLET | January 2004 |
| | | 02248234 | Teva-Lamotrigine 150 MG TABLET | January 2004 |
| | | 02265494 | Mylan-Lamotrigine 25 MG TABLET | April 2005 |
| | | 02265508 | Mylan-Lamotrigine 100 MG TABLET | April 2005 |
| | | 02265516 | Mylan-Lamotrigine 150 MG TABLET | April 2005 |
| | | 02343010 | Lamotrigine 25 MG TABLET | February 2010 |
| | | 02343029 | Lamotrigine 100 MG TABLET | February 2010 |
| | | 02343037 | Lamotrigine 150 MG TABLET | February 2010 |
| | | 02381354 | Auro-Lamotrigine 25 MG TABLET | June 2012 |
| | | 02381362 | Auro-Lamotrigine 100 MG TABLET | June 2012 |
| | | 02381370 | Auro-Lamotrigine 150 MG TABLET | June 2012 |
| | | 02428202 | Lamotrigine 25 MG TABLET | October 2014 |
| | | 02428210 | Lamotrigine 100 MG TABLET | October 2014 |
| | | 02428229 | Lamotrigine 150 MG TABLET | October 2014 |
| | | 02542730 | Jamp Lamotrigine 25 MG TABLET | June 2024 |
| | | 02542749 | Jamp Lamotrigine 100 MG TABLET | June 2024 |
| | | 02542757 | Jamp Lamotrigine 150 MG TABLET | June 2024 |
| Valproic acid | Anticonvulsants (used as mood stabilizer) | 02229628 | Pms-Valproic Acid E.C. 500 MG CAPSULE DR | December 1996 |
| | | 02230768 | Pms-Valproic Acid 250 MG CAPSULE | February 1997 |
| | | 02238048 | Apo-Valproic Acid 250 MG CAPSULE | June 1998 |
| | | 00443832 | Depakene 250 MG/5ML SOLUTION | December 1978 |
| | | 02236807 | Pms-Valproic Acid 250 MG/5ML SOLUTION | November 1997 |
| | | 02238370 | Apo-Valproic Acid Oral Solution 250 MG/5ML SOLUTION | July 1998 |
| | | 02532441 | Jamp Valproic Acid Oral Solution 250 MG/5ML SOLUTION | October 2023 |
| | | 02549190 | Odan-Valproic Acid 250 MG/5ML SOLUTION | November 2024 |
| Risperidone | | 02252007 | Pms-Risperidone 0.25 MG TABLET | July 2006 |

| | | | |
|---|---|---|---|
| | 02252015 | Pms-Risperidone 0.5 MG TABLET | July 2006 |
| | 02252023 | Pms-Risperidone 1 MG TABLET | July 2006 |
| | 02252031 | Pms-Risperidone 2 MG TABLET | July 2006 |
| | 02252058 | Pms-Risperidone 3 MG TABLET | July 2006 |
| | 02252066 | Pms-Risperidone 4 MG TABLET | July 2006 |
| | 02264196 | Teva-Risperidone 1 MG TABLET | July 2006 |
| | 02264218 | Teva-Risperidone 2 MG TABLET | July 2006 |
| | 02264226 | Teva-Risperidone 3 MG TABLET | July 2006 |
| | 02264234 | Teva-Risperidone 4 MG TABLET | July 2006 |
| | 02279266 | Pms-Risperidone 1 MG/ML SOLUTION | July 2006 |
| | 02279800 | Sandoz Risperidone 1 MG TABLET | July 2006 |
| | 02279819 | Sandoz Risperidone 2 MG TABLET | July 2006 |
| 2nd generation (atypical antipsychotics) | 02279827 | Sandoz Risperidone 3 MG TABLET | July 2006 |
| | 02279835 | Sandoz Risperidone 4 MG TABLET | July 2006 |
| | 02282119 | Apo-Risperidone 0.25 MG TABLET | July 2006 |
| | 02282127 | Apo-Risperidone 0.5 MG TABLET | July 2006 |
| | 02282135 | Apo-Risperidone 1 MG TABLET | July 2006 |
| | 02282143 | Apo-Risperidone 2 MG TABLET | July 2006 |
| | 02282151 | Apo-Risperidone 3 MG TABLET | July 2006 |
| | 02282178 | Apo-Risperidone 4 MG TABLET | July 2006 |
| | 02282690 | Teva-Risperidone 0.25 MG TABLET | July 2006 |
| | 02303655 | Sandoz Risperidone 0.25 MG TABLET | December 2007 |
| | 02303663 | Sandoz Risperidone 0.5 MG TABLET | December 2007 |
| | 02328305 | Taro-Risperidone 0.25 MG TABLET | October 2009 |
| | 02328313 | Taro-Risperidone 0.5 MG TABLET | October 2009 |
| | 02328321 | Taro-Risperidone 1 MG TABLET | October 2009 |
| | 02328348 | Taro-Risperidone 2 MG TABLET | October 2009 |
| | 02328364 | Taro-Risperidone 3 MG TABLET | October 2009 |

| | | |
|---|---|---|
| 02328372 | Taro-Risperidone 4 MG TABLET | October 2009 |
| 02332051 | Risperidone Tablets 0.25 MG TABLET | August 2009 |
| 02332078 | Risperidone Tablets 0.5 MG TABLET | August 2009 |
| 02332086 | Risperidone Tablets 1 MG TABLET | August 2009 |
| 02332094 | Risperidone Tablets 2 MG TABLET | August 2009 |
| 02332108 | Risperidone Tablets 3 MG TABLET | August 2009 |
| 02332116 | Risperidone Tablets 4 MG TABLET | August 2009 |
| 02356880 | Risperidone 0.25 MG TABLET | October 2010 |
| 02356899 | Risperidone 0.5 MG TABLET | October 2010 |
| 02356902 | Risperidone 1 MG TABLET | October 2010 |
| 02356910 | Risperidone 2 MG TABLET | October 2010 |
| 02356929 | Risperidone 3 MG TABLET | October 2010 |
| 02356937 | Risperidone 4 MG TABLET | October 2010 |
| 02359529 | Jamp-Risperidone 0.25 MG TABLET | December 2010 |
| 02359537 | Jamp-Risperidone 0.5 MG TABLET | December 2010 |
| 02359545 | Jamp-Risperidone 1 MG TABLET | December 2010 |
| 02359553 | Jamp-Risperidone 2 MG TABLET | December 2010 |
| 02359561 | Jamp-Risperidone 3 MG TABLET | December 2010 |
| 02359588 | Jamp-Risperidone 4 MG TABLET | December 2010 |
| 02359790 | Mint-Risperidon 0.25 MG TABLET | January 2011 |
| 02359804 | Mint-Risperidon 0.5 MG TABLET | January 2011 |
| 02359812 | Mint-Risperidon 1 MG TABLET | January 2011 |
| 02359820 | Mint-Risperidon 2 MG TABLET | January 2011 |
| 02359839 | Mint-Risperidon 3 MG TABLET | January 2011 |
| 02359847 | Mint-Risperidon 4 MG TABLET | January 2011 |
| 02371766 | Mar-Risperidone 0.25 MG TABLET | April 2013 |
| 02371774 | Mar-Risperidone 0.5 MG TABLET | April 2013 |
| 02371782 | Mar-Risperidone 1 MG TABLET | April 2013 |

| | | | | |
|---|---|---|---|---|
| | | 02454319 | Jamp-Risperidone 1 MG/ML SOLUTION | August 2018 |
| | | 02533804 | Risperidone 0.25 MG TABLET | November 2023 |
| | | 02533928 | Risperidone 0.5 MG TABLET | November 2023 |
| | | 02533936 | Risperidone 1 MG TABLET | November 2023 |
| | | 02533944 | Risperidone 2 MG TABLET | November 2023 |
| | | 02533952 | Risperidone 3 MG TABLET | November 2023 |
| | | 02533960 | Risperidone 4 MG TABLET | November 2023 |
| | | 02255707 | Risperdal Consta 25 MG/2 ML VIAL | November 2004 |
| | | 02255723 | Risperdal Consta 37.5MG/2ML VIAL | November 2004 |
| | | 02255758 | Risperdal Consta 50 MG/2 ML VIAL | November 2004 |
| | | 02298465 | Risperdal Consta 12.5MG/2ML VIAL | October 2010 |
| Olanzapine | 2nd generation (atypical antipsychotics) | 02229250 | Zyprexa 2.5 MG TABLET | December 1997 |
| | | 02229269 | Zyprexa 5 MG TABLET | November 1996 |
| | | 02229277 | Zyprexa 7.5 MG TABLET | October 1996 |
| | | 02229285 | Zyprexa 10 MG TABLET | November 1996 |
| | | 02238850 | Zyprexa 15 MG TABLET | June 2002 |
| | | 02238851 | Zyprexa 20 MG TABLET | April 2007 |
| | | 02243086 | Zyprexa Zydis 5 MG TAB RAPDIS | March 2001 |
| | | 02243087 | Zyprexa Zydis 10 MG TAB RAPDIS | March 2001 |
| | | 02243088 | Zyprexa Zydis 15 MG TAB RAPDIS | March 2001 |
| | | 02243089 | Zyprexa Zydis 20 MG TAB RAPDIS | March 2001 |
| | | 02276712 | Teva-Olanzapine 2.5 MG TABLET | June 2007 |
| | | 02276720 | Teva-Olanzapine 5 MG TABLET | June 2007 |
| | | 02276739 | Teva-Olanzapine 7.5 MG TABLET | June 2007 |
| | | 02276747 | Teva-Olanzapine 10 MG TABLET | June 2007 |
| | | 02276755 | Teva-Olanzapine 15 MG TABLET | June 2007 |
| | | 02281791 | Apo-Olanzapine 2.5 MG TABLET | October 2009 |
| | | 02281805 | Apo-Olanzapine 5 MG TABLET | October 2009 |

| | | |
|---|---|---|
| 02281813 | Apo-Olanzapine 7.5 MG TABLET | October 2009 |
| 02281821 | Apo-Olanzapine 10 MG TABLET | October 2009 |
| 02281848 | Apo-Olanzapine 15 MG TABLET | October 2009 |
| 02303116 | Pms-Olanzapine 2.5 MG TABLET | April 2008 |
| 02303159 | Pms-Olanzapine 5 MG TABLET | April 2008 |
| 02303167 | Pms-Olanzapine 7.5 MG TABLET | April 2008 |
| 02303175 | Pms-Olanzapine 10 MG TABLET | April 2008 |
| 02303183 | Pms-Olanzapine 15 MG TABLET | April 2008 |
| 02303191 | Pms-Olanzapine Odt 5 MG TAB RAPDIS | October 2009 |
| 02303205 | Pms-Olanzapine Odt 10 MG TAB RAPDIS | October 2009 |
| 02303213 | Pms-Olanzapine Odt 15 MG TAB RAPDIS | October 2009 |
| 02307391 | Dom-Olanzapine Odt 15 MG TAB RAPDIS | October 2009 |
| 02310341 | Sandoz Olanzapine 2.5 MG TABLET | December 2009 |
| 02310368 | Sandoz Olanzapine 5 MG TABLET | December 2009 |
| 02310376 | Sandoz Olanzapine 7.5 MG TABLET | December 2009 |
| 02310384 | Sandoz Olanzapine 10 MG TABLET | December 2009 |
| 02310392 | Sandoz Olanzapine 15 MG TABLET | December 2009 |
| 02311968 | Olanzapine 2.5 MG TABLET | February 2010 |
| 02311976 | Olanzapine 5 MG TABLET | February 2010 |
| 02311984 | Olanzapine 7.5 MG TABLET | February 2010 |
| 02311992 | Olanzapine 10 MG TABLET | February 2010 |
| 02312018 | Olanzapine 15 MG TABLET | February 2010 |
| 02327775 | Sandoz Olanzapine Odt 5 MG TAB RAPDIS | February 2010 |
| 02327783 | Sandoz Olanzapine Odt 10 MG TAB RAPDIS | February 2010 |
| 02327791 | Sandoz Olanzapine Odt 15 MG TAB RAPDIS | February 2010 |
| 02327805 | Sandoz Olanzapine Odt 20 MG TAB RAPDIS | February 2010 |
| 02333015 | Apo-Olanzapine 20 MG TABLET | October 2009 |
| 02337169 | Riva-Olanzapine 15 MG TABLET | February 2010 |

| | | |
|---|---|---|
| 02338645 | Olanzapine Odt 5 MG TAB RAPDIS | February 2010 |
| 02338653 | Olanzapine Odt 10 MG TAB RAPDIS | February 2010 |
| 02338661 | Olanzapine Odt 15 MG TAB RAPDIS | February 2010 |
| 02343665 | Olanzapine Odt 5 MG TAB RAPDIS | April 2012 |
| 02343673 | Olanzapine Odt 10 MG TAB RAPDIS | April 2012 |
| 02343681 | Olanzapine Odt 15 MG TAB RAPDIS | April 2012 |
| 02343703 | Olanzapine Odt 20 MG TAB RAPDIS | April 2012 |
| 02352974 | Olanzapine Odt 5 MG TAB RAPDIS | October 2011 |
| 02352982 | Olanzapine Odt 10 MG TAB RAPDIS | October 2011 |
| 02352990 | Olanzapine Odt 15 MG TAB RAPDIS | October 2011 |
| 02359707 | Teva-Olanzapine 20 MG TABLET | April 2011 |
| 02360616 | Apo-Olanzapine Odt 5 MG TAB RAPDIS | November 2011 |
| 02360624 | Apo-Olanzapine Odt 10 MG TAB RAPDIS | August 2011 |
| 02360632 | Apo-Olanzapine Odt 15 MG TAB RAPDIS | August 2011 |
| 02360640 | Apo-Olanzapine Odt 20 MG TAB RAPDIS | August 2011 |
| 02372819 | Olanzapine 2.5 MG TABLET | October 2011 |
| 02372827 | Olanzapine 5 MG TABLET | October 2011 |
| 02372835 | Olanzapine 7.5 MG TABLET | October 2011 |
| 02372843 | Olanzapine 10 MG TABLET | October 2011 |
| 02372851 | Olanzapine 15 MG TABLET | October 2011 |
| 02385864 | Olanzapine 2.5 MG TABLET | June 2012 |
| 02385872 | Olanzapine 5 MG TABLET | June 2012 |
| 02385880 | Olanzapine 7.5 MG TABLET | June 2012 |
| 02385899 | Olanzapine 10 MG TABLET | June 2012 |
| 02385902 | Olanzapine 15 MG TABLET | June 2012 |
| 02385910 | Olanzapine 20 MG TABLET | July 2023 |
| 02406624 | Jamp Olanzapine Odt 5 MG TAB RAPDIS | August 2014 |
| 02406632 | Jamp Olanzapine Odt 10 MG TAB RAPDIS | August 2014 |

| | | |
|---|---|---|
| 02406640 | Jamp Olanzapine Odt 15 MG TAB RAPDIS | August 2014 |
| 02406659 | Jamp Olanzapine Odt 20 MG TAB RAPDIS | August 2014 |
| 02410141 | Mint-Olanzapine 2.5 MG TABLET | September 2019 |
| 02410168 | Mint-Olanzapine 5 MG TABLET | September 2019 |
| 02410176 | Mint-Olanzapine 7.5 MG TABLET | September 2019 |
| 02410184 | Mint-Olanzapine 10 MG TABLET | September 2019 |
| 02410192 | Mint-Olanzapine 15 MG TABLET | September 2019 |
| 02417243 | Jamp Olanzapine Fc 2.5 MG TABLET | June 2015 |
| 02417251 | Jamp Olanzapine Fc 5 MG TABLET | June 2015 |
| 02417278 | Jamp Olanzapine Fc 7.5 MG TABLET | June 2015 |
| 02417286 | Jamp Olanzapine Fc 10 MG TABLET | June 2015 |
| 02417294 | Jamp Olanzapine Fc 15 MG TABLET | June 2015 |
| 02417308 | Jamp Olanzapine Fc 20 MG TABLET | June 2015 |
| 02421704 | Olanzapine 20 MG TABLET | June 2014 |
| 02425114 | Olanzapine Odt 20 MG TAB RAPDIS | June 2014 |
| 02436965 | Mint-Olanzapine Odt 5 MG TAB RAPDIS | March 2015 |
| 02436973 | Mint-Olanzapine Odt 10 MG TAB RAPDIS | March 2015 |
| 02436981 | Mint-Olanzapine Odt 15 MG TAB RAPDIS | March 2015 |
| 02448726 | Auro-Olanzapine Odt 5 MG TAB RAPDIS | December 2015 |
| 02448734 | Auro-Olanzapine Odt 10 MG TAB RAPDIS | December 2015 |
| 02448742 | Auro-Olanzapine Odt 15 MG TAB RAPDIS | December 2015 |
| 02448750 | Auro-Olanzapine Odt 20 MG TAB RAPDIS | December 2015 |
| 02536188 | Nra-Olanzapine Odt 5 MG TAB RAPDIS | April 2024 |
| 02536196 | Nra-Olanzapine Odt 10 MG TAB RAPDIS | April 2024 |
| 02536218 | Nra-Olanzapine Odt 15 MG TAB RAPDIS | April 2024 |
| 02536226 | Nra-Olanzapine Odt 20 MG TAB RAPDIS | April 2024 |
| 02545586 | Nra-Olanzapine 2.5 MG TABLET | April 2024 |
| 02545594 | Nra-Olanzapine 5 MG TABLET | April 2024 |

| | | | | |
|---|---|---|---|---|
| | | 02545608 | Nra-Olanzapine 7.5 MG TABLET | April 2024 |
| | | 02545616 | Nra-Olanzapine 10 MG TABLET | April 2024 |
| | | 02545624 | Nra-Olanzapine 15 MG TABLET | April 2024 |
| | | 02545632 | Nra-Olanzapine 20 MG TABLET | April 2024 |
| Aripiprazole | 2nd generation (atypical antipsychotics) | 02322374 | Abilify 2 MG TABLET | September 2009 |
| | | 02322382 | Abilify 5 MG TABLET | September 2009 |
| | | 02322390 | Abilify 10 MG TABLET | September 2009 |
| | | 02322404 | Abilify 15 MG TABLET | September 2009 |
| | | 02322412 | Abilify 20 MG TABLET | September 2009 |
| | | 02322455 | Abilify 30 MG TABLET | September 2009 |
| | | 02420864 | Abilify Maintena 300 MG SUSER VIAL | March 2014 |
| | | 02420872 | Abilify Maintena 400 MG SUSER VIAL | March 2014 |
| | | 02460025 | Auro-Aripiprazole 2 MG TABLET | July 2018 |
| | | 02460033 | Auro-Aripiprazole 5 MG TABLET | July 2018 |
| | | 02460041 | Auro-Aripiprazole 10 MG TABLET | July 2018 |
| | | 02460068 | Auro-Aripiprazole 15 MG TABLET | July 2018 |
| | | 02460076 | Auro-Aripiprazole 20 MG TABLET | July 2018 |
| | | 02460084 | Auro-Aripiprazole 30 MG TABLET | July 2018 |
| | | 02466635 | Pms-Aripiprazole 2 MG TABLET | July 2018 |
| | | 02466643 | Pms-Aripiprazole 5 MG TABLET | July 2018 |
| | | 02466651 | Pms-Aripiprazole 10 MG TABLET | July 2018 |
| | | 02466678 | Pms-Aripiprazole 15 MG TABLET | July 2018 |
| | | 02466686 | Pms-Aripiprazole 20 MG TABLET | July 2018 |
| | | 02466694 | Pms-Aripiprazole 30 MG TABLET | July 2018 |
| | | 02471086 | Apo-Aripiprazole 2 MG TABLET | April 2018 |
| | | 02471094 | Apo-Aripiprazole 5 MG TABLET | April 2018 |
| | | 02471108 | Apo-Aripiprazole 10 MG TABLET | April 2018 |
| | | 02471116 | Apo-Aripiprazole 15 MG TABLET | April 2018 |

| | | |
|---|---|---|
| 02471124 | Apo-Aripiprazole 20 MG TABLET | April 2018 |
| 02471132 | Apo-Aripiprazole 30 MG TABLET | April 2018 |
| 02472201 | Nra-Aripiprazole 2 MG TABLET | January 2024 |
| 02472228 | Nra-Aripiprazole 5 MG TABLET | January 2024 |
| 02472244 | Nra-Aripiprazole 10 MG TABLET | January 2024 |
| 02472252 | Nra-Aripiprazole 15 MG TABLET | January 2024 |
| 02472260 | Nra-Aripiprazole 20 MG TABLET | January 2024 |
| 02472279 | Nra-Aripiprazole 30 MG TABLET | January 2024 |
| 02473658 | Sandoz Aripiprazole 2 MG TABLET | July 2018 |
| 02473666 | Sandoz Aripiprazole 5 MG TABLET | July 2018 |
| 02473674 | Sandoz Aripiprazole 10 MG TABLET | July 2018 |
| 02473682 | Sandoz Aripiprazole 15 MG TABLET | July 2018 |
| 02473690 | Sandoz Aripiprazole 20 MG TABLET | July 2018 |
| 02473704 | Sandoz Aripiprazole 30 MG TABLET | July 2018 |
| 02483556 | Mint-Aripiprazole 2 MG TABLET | May 2020 |
| 02483564 | Mint-Aripiprazole 5 MG TABLET | April 2020 |
| 02483572 | Mint-Aripiprazole 10 MG TABLET | April 2020 |
| 02483580 | Mint-Aripiprazole 15 MG TABLET | April 2020 |
| 02483599 | Mint-Aripiprazole 20 MG TABLET | April 2020 |
| 02483602 | Mint-Aripiprazole 30 MG TABLET | April 2020 |
| 02506688 | Aripiprazole 2 MG TABLET | April 2021 |
| 02506718 | Aripiprazole 5 MG TABLET | April 2021 |
| 02506726 | Aripiprazole 10 MG TABLET | April 2021 |
| 02506734 | Aripiprazole 15 MG TABLET | April 2021 |
| 02506750 | Aripiprazole 20 MG TABLET | April 2021 |
| 02506785 | Aripiprazole 30 MG TABLET | April 2021 |
| 02534320 | Aripiprazole 2 MG TABLET | July 2023 |
| 02534339 | Aripiprazole 5 MG TABLET | July 2023 |

| | | | | |
|---|---|---|---|---|
| | | 02534347 | Aripiprazole 10 MG TABLET | July 2023 |
| | | 02534355 | Aripiprazole 15 MG TABLET | July 2023 |
| | | 02534363 | Aripiprazole 20 MG TABLET | July 2023 |
| | | 02534371 | Aripiprazole 30 MG TABLET | July 2023 |
| | | 02553163 | Jamp Aripiprazole 2 MG TABLET | November 2025 |
| | | 02553171 | Jamp Aripiprazole 5 MG TABLET | November 2025 |
| | | 02553198 | Jamp Aripiprazole 10 MG TABLET | November 2025 |
| | | 02553201 | Jamp Aripiprazole 15 MG TABLET | November 2025 |
| | | 02553228 | Jamp Aripiprazole 20 MG TABLET | November 2025 |
| | | 02553236 | Jamp Aripiprazole 30 MG TABLET | November 2025 |
| | | 02554569 | Abilify Asimtufii 720 MG/2.4 SUSER SYR | March 2025 |
| | | 02554577 | Abilify Asimtufii 960 MG/3.2 SUSER SYR | March 2025 |
| Quetiapine | 2nd generation (atypical antipsychotics) | 02236951 | Seroquel 25 MG TABLET | December 1997 |
| | | 02236952 | Seroquel 100 MG TABLET | December 1997 |
| | | 02236953 | Seroquel 200 MG TABLET | December 1997 |
| | | 02244107 | Seroquel 300 MG TABLET | August 2001 |
| | | 02296551 | Pms-Quetiapine 25 MG TABLET | September 2008 |
| | | 02296578 | Pms-Quetiapine 100 MG TABLET | September 2008 |
| | | 02296594 | Pms-Quetiapine 200 MG TABLET | September 2008 |
| | | 02296608 | Pms-Quetiapine 300 MG TABLET | September 2008 |
| | | 02300184 | Seroquel Xr 50 MG TAB ER 24H | September 2007 |
| | | 02300192 | Seroquel Xr 200 MG TAB ER 24H | September 2007 |
| | | 02300206 | Seroquel Xr 300 MG TAB ER 24H | September 2007 |
| | | 02300214 | Seroquel Xr 400 MG TAB ER 24H | September 2007 |
| | | 02316080 | Act Quetiapine 25 MG TABLET | September 2008 |
| | | 02316099 | Act Quetiapine 100 MG TABLET | September 2008 |
| | | 02316110 | Act Quetiapine 200 MG TABLET | September 2008 |
| | | 02316129 | Act Quetiapine 300 MG TABLET | September 2008 |

| | | |
|---|---|---|
| 02316692 | Riva-Quetiapine 25 MG TABLET | September 2008 |
| 02316706 | Riva-Quetiapine 100 MG TABLET | September 2008 |
| 02316722 | Riva-Quetiapine 200 MG TABLET | September 2008 |
| 02316730 | Riva-Quetiapine 300 MG TABLET | September 2008 |
| 02317346 | Pro-Quetiapine 25 MG TABLET | October 2008 |
| 02317354 | Pro-Quetiapine 100 MG TABLET | October 2008 |
| 02317362 | Pro-Quetiapine 200 MG TABLET | October 2008 |
| 02317370 | Pro-Quetiapine 300 MG TABLET | October 2008 |
| 02317893 | Quetiapine 25 MG TABLET | February 2009 |
| 02317907 | Quetiapine 100 MG TABLET | February 2009 |
| 02317923 | Quetiapine 200 MG TABLET | February 2009 |
| 02317931 | Quetiapine 300 MG TABLET | February 2009 |
| 02321513 | Seroquel Xr 150 MG TAB ER 24H | April 2009 |
| 02330423 | Jamp-Quetiapine 100 MG TABLET | April 2010 |
| 02353164 | Quetiapine 25 MG TABLET | July 2010 |
| 02353172 | Quetiapine 100 MG TABLET | July 2010 |
| 02353199 | Quetiapine 200 MG TABLET | July 2010 |
| 02353202 | Quetiapine 300 MG TABLET | July 2010 |
| 02361892 | Pms-Quetiapine 50 MG TABLET | January 2011 |
| 02387794 | Ach-Quetiapine 25 MG TABLET | June 2012 |
| 02387808 | Ach-Quetiapine 100 MG TABLET | June 2012 |
| 02387824 | Ach-Quetiapine 200 MG TABLET | June 2012 |
| 02387832 | Ach-Quetiapine 300 MG TABLET | June 2012 |
| 02390140 | Jamp Quetiapine Fumarate 25 MG TABLET | November 2021 |
| 02390159 | Jamp Quetiapine Fumarate 100 MG TABLET | November 2021 |
| 02390167 | Jamp Quetiapine Fumarate 200 MG TABLET | November 2021 |
| 02390175 | Jamp Quetiapine Fumarate 300 MG TABLET | November 2021 |
| 02390205 | Auro-Quetiapine 25 MG TABLET | October 2012 |

| | | |
|---|---|---|
| 02390213 | Auro-Quetiapine 100 MG TABLET | October 2012 |
| 02390248 | Auro-Quetiapine 200 MG TABLET | October 2012 |
| 02390256 | Auro-Quetiapine 300 MG TABLET | October 2012 |
| 02395444 | Teva-Quetiapine Xr 50 MG TAB ER 24H | March 2013 |
| 02395452 | Teva-Quetiapine Xr 150 MG TAB ER 24H | March 2013 |
| 02395460 | Teva-Quetiapine Xr 200 MG TAB ER 24H | March 2013 |
| 02395479 | Teva-Quetiapine Xr 300 MG TAB ER 24H | March 2013 |
| 02395487 | Teva-Quetiapine Xr 400 MG TAB ER 24H | March 2013 |
| 02407671 | Sandoz Quetiapine Xrt 50 MG TAB ER 24H | June 2013 |
| 02407698 | Sandoz Quetiapine Xrt 150 MG TAB ER 24H | June 2013 |
| 02407701 | Sandoz Quetiapine Xrt 200 MG TAB ER 24H | June 2013 |
| 02407728 | Sandoz Quetiapine Xrt 300 MG TAB ER 24H | June 2013 |
| 02407736 | Sandoz Quetiapine Xrt 400 MG TAB ER 24H | June 2013 |
| 02417359 | Quetiapine Xr 50 MG TAB ER 24H | January 2014 |
| 02417367 | Quetiapine Xr 150 MG TAB ER 24H | January 2014 |
| 02417375 | Quetiapine Xr 200 MG TAB ER 24H | January 2014 |
| 02417383 | Quetiapine Xr 300 MG TAB ER 24H | January 2014 |
| 02417391 | Quetiapine Xr 400 MG TAB ER 24H | January 2014 |
| 02438003 | Mint-Quetiapine 25 MG TABLET | March 2015 |
| 02438011 | Mint-Quetiapine 100 MG TABLET | March 2015 |
| 02438046 | Mint-Quetiapine 200 MG TABLET | March 2015 |
| 02438054 | Mint-Quetiapine 300 MG TABLET | March 2015 |
| 02439158 | Nat-Quetiapine 25 MG TABLET | July 2015 |
| 02439166 | Nat-Quetiapine 100 MG TABLET | July 2015 |
| 02439182 | Nat-Quetiapine 200 MG TABLET | July 2015 |
| 02439190 | Nat-Quetiapine 300 MG TABLET | July 2015 |
| 02447193 | Bio-Quetiapine 25 MG TABLET | September 2016 |
| 02450860 | Ach-Quetiapine Fumarate Xr 50 MG TAB ER 24H | October 2020 |

| | | |
|---|---|---|
| 02450879 | Ach-Quetiapine Fumarate Xr 150 MG TAB ER 24H | October 2020 |
| 02450887 | Ach-Quetiapine Fumarate Xr 200 MG TAB ER 24H | October 2020 |
| 02450895 | Ach-Quetiapine Fumarate Xr 300 MG TAB ER 24H | January 2021 |
| 02450909 | Ach-Quetiapine Fumarate Xr 400 MG TAB ER 24H | October 2020 |
| 02457229 | Apo-Quetiapine Xr 50 MG TAB ER 24H | June 2018 |
| 02457237 | Apo-Quetiapine Xr 150 MG TAB ER 24H | June 2018 |
| 02457245 | Apo-Quetiapine Xr 200 MG TAB ER 24H | June 2018 |
| 02457253 | Apo-Quetiapine Xr 300 MG TAB ER 24H | June 2018 |
| 02457261 | Apo-Quetiapine Xr 400 MG TAB ER 24H | June 2018 |
| 02486237 | Nra-Quetiapine 25 MG TABLET | September 2019 |
| 02501635 | Apo-Quetiapine Fumarate 25 MG TABLET | July 2021 |
| 02501643 | Apo-Quetiapine Fumarate 100 MG TABLET | July 2021 |
| 02501651 | Apo-Quetiapine Fumarate 200 MG TABLET | July 2021 |
| 02501678 | Apo-Quetiapine Fumarate 300 MG TABLET | July 2021 |
| 02510677 | Nra-Quetiapine Xr 50 MG TAB ER 24H | July 2021 |
| 02510685 | Nra-Quetiapine Xr 150 MG TAB ER 24H | July 2021 |
| 02510693 | Nra-Quetiapine Xr 200 MG TAB ER 24H | July 2021 |
| 02510707 | Nra-Quetiapine Xr 300 MG TAB ER 24H | July 2021 |
| 02510715 | Nra-Quetiapine Xr 400 MG TAB ER 24H | July 2021 |
| 02516616 | Quetiapine Fumarate Xr 50 MG TAB ER 24H | December 2021 |
| 02516624 | Quetiapine Fumarate Xr 150 MG TAB ER 24H | December 2021 |
| 02516632 | Quetiapine Fumarate Xr 200 MG TAB ER 24H | December 2021 |
| 02516640 | Quetiapine Fumarate Xr 300 MG TAB ER 24H | December 2021 |
| 02516659 | Quetiapine Fumarate Xr 400 MG TAB ER 24H | December 2021 |
| 02519607 | Quetiapine Xr 50 MG TAB ER 24H | April 2022 |
| 02519615 | Quetiapine Xr 150 MG TAB ER 24H | April 2022 |
| 02519623 | Quetiapine Xr 200 MG TAB ER 24H | April 2022 |
| 02519747 | Quetiapine Xr 300 MG TAB ER 24H | April 2022 |

| | | |
|---|---|---|
| 02519763 | Quetiapine Xr 400 MG TAB ER 24H | April 2022 |
| 02522187 | Mint-Quetiapine Xr 50 MG TAB ER 24H | April 2022 |
| 02522195 | Mint-Quetiapine Xr 150 MG TAB ER 24H | April 2022 |
| 02522209 | Mint-Quetiapine Xr 200 MG TAB ER 24H | April 2022 |
| 02522217 | Mint-Quetiapine Xr 300 MG TAB ER 24H | April 2022 |
| 02522225 | Mint-Quetiapine Xr 400 MG TAB ER 24H | October 2022 |
| 02527928 | M-Quetiapine Fumarate Xr 50 MG TAB ER 24H | March 2023 |
| 02527936 | M-Quetiapine Fumarate Xr 150 MG TAB ER 24H | March 2023 |
| 02527944 | M-Quetiapine Fumarate Xr 200 MG TAB ER 24H | March 2023 |
| 02527952 | M-Quetiapine Fumarate Xr 300 MG TAB ER 24H | March 2023 |
| 02527960 | M-Quetiapine Fumarate Xr 400 MG TAB ER 24H | March 2023 |

Abbreviations – DIN/PIN: Drug identification number/Product identification number.

[1] Retrieved from https://pharmacareformularysearch.gov.bc.ca/SearchResults.xhtml; https://www.formulary.health.gov.on.ca/formulary/

[2] Retrieved from https://health-products.canada.ca/dpd-bdpp/?lang=eng

**Appendix Table 5.** Treatments options for bipolar disorder at recommended dosage (mg/day).

| Treatment | Week 1 | Week 2 | Week 3 | Week 4 and beyond |
|---|---|---|---|---|
| Lithium[117–119] | ≤300 | 300-600 | 600-900 | 900-1200 |
| Divalproex[120] | 500-750 | 750-1500 | 1000-2000 | 1500-2000 |
| Lamotrigine[121,122] | 25 | 25 | 50 | 50-200 |
| Valproic acid[120] | 500-750 | 750-1500 | 1000-2000 | 1500-2500 |
| Risperidone[123,124] | 1-3 | 2-3 | 3-4 | 4-6 |
| Olanzapine[125] | 5-20 | 10-20 | 10-20 | 10-20 |
| Aripiprazole[126–128] | 10-15 | 15 | 20-30 | 15-30 |
| Quetiapine[129] | 300-600 | 300-900 | 300-900 | 600-900 |

21. American Psychiatric Association. *Practice guideline for the treatment of patients with bipolar disorder (revision)*. American Psychiatric Pub, 2002.

31. Bowden CL, Calabrese JR, McElroy SL, *et al.* A randomized, placebo-controlled 12-month

trial of divalproex and lithium in treatment of outpatients with bipolar I disorder. *Arch Gen Psychiatry* 2000; **57**: 481–489.

40. Cipriani A, Reid K, Young AH, Macritchie K, Geddes J. Valproic acid, valproate and divalproex in the maintenance treatment of bipolar disorder. *Cochrane Database Syst Rev* 2013.

58. CAMH Bipolar Clinic Staff. *Bipolar disorder: An information guide (revised edition).*, 2013.

59. Ehmann T, Hanson L. *Early Psychosis: A Care Guide*. Vancouver, BC, 2022.

68. Buckley P, Thompson P, Way L, Meltzer HY. Substance abuse among patients with treatment-resistant schizophrenia: characteristics and implications for clozapine therapy. *Am J Psychiatry* 1994.

69. McEvoy J, Freudenreich O, McGee M, Vanderzwaag C, Levin E, Rose J. Clozapine decreases smoking in patients with chronic schizophrenia. *Biol Psychiatry* 1995; **37**: 550–552.

70. Drake RE, Xie H, McHugo GJ, Green AI. The effects of clozapine on alcohol and drug use disorders among patients with schizophrenia. *Schizophr Bull* 2000; **26**: 441–449.

71. Zimmet S V, Strous RD, Burgess ES, Kohnstamm S, Green AI. Effects of clozapine on substance use in patients with schizophrenia and schizoaffective disorder: a retrospective survey. *J Clin Psychopharmacol* 2000; **20**: 94–98.

72. Anderson E, Dive L, Kang S, Newth S, Vidal C, Bilsker D. *Family physician guide for depression, anxiety disorders, early psychosis and substance use disorders*. Burnaby, BC, 2007.

73. Canadian Network for Mood and Anxiety Treatments (CANMAT). The treatment of bipolar disorder: review of the literature, guidelines, and options. *Can J Psychiatry* 1997; **42**: 67S–100S.

74. Goodwin GM. Evidence-based guidelines for treating bipolar disorder: recommendations from the British Association for Psychopharmacology. *J Psychopharmacol* 2003; **17**: 149–173.

75. BC Reproductive Mental Health Program & Perinatal Services BC. *Best Practice Guidelines for Mental Health Disorders in the Perinatal Period*., 2014.

84. National Institute for Health and Care Excellence. *Bipolar disorder: assessment and management*. London: NICE, National Institute for Health and Care Excellence, 2023.

104. Oquendo MA, Galfalvy HC, Currier D, *et al.* Treatment of suicide attempters with bipolar disorder: a randomized clinical trial comparing lithium and valproate in the prevention of

suicidal behavior. *Am J Psychiatry* 2011; **168**: 1050–1056.

120. Macaluso M, Johnston C. *Handbook of Practical Psychopharmacology*. 1st ed. Washington DC: John Wiley & Sons, 2025.

121. Hashimoto Y, Kotake K, Watanabe N, Fujiwara T, Sakamoto S. Lamotrigine in the maintenance treatment of bipolar disorder. *Cochrane Database Syst Rev* 2021.
